# Mental disorders, mortality following myocardial infarction and the impact of the COVID-19 pandemic in England: a cohort study

**DOI:** 10.1101/2025.11.21.25340653

**Authors:** Kelly Fleetwood, John Nolan, Colin Berry, Debbie Cavers, Stewart W Mercer, Sandosh Padmanabhan, Daniel J Smith, Robert Stewart, Amanda Vettini, Caroline A Jackson, the CVD-COVID-UK/COVID-IMPACT Consortium

**Affiliations:** Usher Institute, University of Edinburgh, Edinburgh, UK; British Heart Foundation Data Science Centre, Health Data Research UK, London, UK; School of Cardiovascular and Metabolic Health, University of Glasgow, Glasgow, UK; Centre for Clinical Brain Sciences, University of Edinburgh, UK; Department of Psychological Medicine, King’s College London, London, UK; South London and Maudsley NHS Foundation Trust, London, UK

## Abstract

**Background and Aims:** People with schizophrenia experience poorer cardiovascular disease (CVD) outcomes, but other mental disorders have been little studied. We compared post-myocardial infarction (MI) mortality by mental disorder, explored whether differences in care contributed to mortality disparities, and investigated whether disparities worsened during the COVID-19 pandemic.

**Methods:** We identified people with MI in England (November 2019 – February 2023) from the Myocardial Ischaemia National Audit Programme, ascertaining mental disorder diagnoses from linked electronic health records and mortality from national death records. We used logistic regression to compare 30-day and one-year mortality between people with each of schizophrenia, bipolar disorder or depression (of any severity) versus those without these disorders for ST-elevation MI (STEMI) and non-STEMI (NSTEMI), adjusting for confounders and investigating differences by calendar time period. For one-year mortality, we additionally adjusted for receipt of guideline-informed care.

**Results:** We included 131,075 patients with NSTEMI and 79,045 with STEMI. For NSTEMI, schizophrenia [odds ratio (OR) 1.73, 95% CI 1.20–2.49] and depression (1.17, 1.08–1.26) were associated with higher 30-day mortality. For STEMI, 30-day mortality was higher in people with schizophrenia (1.61, 1.09–2.37), bipolar disorder (1.68, 1.08–2.59) and depression (1.10, 1.01–1.20). All disorders were associated with higher one-year mortality following NSTEMI and STEMI, with adjustment for care attenuating estimates. Associations were generally unaffected by the COVID-19 pandemic.

**Conclusions:** Our findings highlight the increased risk of post-MI mortality in people with mental disorders and support the urgent need to improve implementation of acute cardiac care standards in this group.

**Summary:** We used linked electronic health records from over 200 000 NSTEMI and STEMI patients in England to compare mortality following myocardial infarction by mental disorder status, evaluate whether differences in receipt of acute MI care by mental disorder might help to explain any disparities in mortality, and assess how any disparities in mortality changed following the COVID-19 pandemic. We identified mortality disparities for both NSTEMI and STEMI, which were greatest for people with schizophrenia, but also evident for bipolar disorder and depression. Our findings suggest that differences in receipt of care contributed to poorer survival at one year. Disparities were generally unaffected by the COVID-19 pandemic.

## Introduction

Mental disorders are associated with an elevated risk of, and poor outcomes from, cardiovascular disease.^1,2^ Studies from both universal and non-universal healthcare settings have demonstrated that following myocardial infarction (MI), people with mental disorders have a higher 30-day and one-year mortality^3^, a disparity that persists in recent decades.^4,5^ Previous studies have largely focused on schizophrenia^5–10^, with little investigation of other conditions such as bipolar disorder and depression.^4,11,12^ Analysis by type of MI is scarce and limited to non-universal healthcare settings.^13–15^ Given differences in clinical management and survival of NSTEMI and STEMI, we might anticipate mental disorder-related outcome disparities to differ by MI type. Whilst the mechanisms underlying the observed MI mortality disparities remain poorly understood, these are likely multifactorial^2^ and may include differences in receipt of acute MI care, with studies consistently finding a lower receipt of percutaneous coronary intervention (PCI) in people with a mental disorder.^3^ We recently found that, in England, people diagnosed with a mental disorder had lower receipt of clinical guideline-informed care standards for NSTEMI, including receipt and timeliness of angiogram, admission to cardiac ward, receipt of secondary prevention medication prescribing on discharge and cardiac rehabilitation referral.^16^ We also found some differences in receipt of care standards for STEMI, such as timeliness of revascularisation, although patterns varied by mental disorder and care standard. However, the contribution of disparities in clinical care to MI survival by mental disorder status remain little investigated, with inconsistent findings from studies that adjusted mental disorder-mortality associations for receipt of revascularisation.^3,7,17^

Furthermore, the response to the COVID-19 pandemic led to significant health care delivery disruption, including reduced cardiovascular care activity.^18–20^ Studies also revealed greater delays from symptom onset to seeking medical help for ischaemic heart disease (IHD) and increased short-term mortality during the early period of the pandemic compared to the pre-pandemic period.^21^ To our knowledge, no study has examined whether the pandemic exacerbated existing mental disorder disparities in survival following MI.

In this study, for each of NSTEMI and STEMI, we sought to: i) compare 30-day and 1-year mortality following MI among people with versus without schizophrenia, bipolar disorder or depression of any severity; ii) explore the extent to which disparities in receipt of care might explain disparities in mortality; and iii) determine whether mortality disparities worsened during the COVID-19 pandemic.

## Methods

We have reported this study in accordance with the Reporting of Studies Conducted using Observational Routinely-Collected Health Data (RECORD) statement.^22^

### Setting and data sources

We accessed national linked electronic health records from England in NHS England’s (NHSE) Secure Data Environment (SDE), made available through the British Heart Foundation Data Science Centre’s <u>CVD-COVID-UK/COVID-IMPACT Consortium</u>.^23^ Our primary data source was the Myocardial Ischaemia National Audit Project (MINAP), a clinical audit of patients admitted to hospital with MI, which includes information on patient demographics, medical history, clinical characteristics of the MI and care from the call for help to discharge.^24^ We also used data from the General Practice Extraction Service (GPES) Data for Pandemic Planning and Research (GDPPR) dataset which captures data from 98% of GP practices in England, and includes diagnoses from the primary care domain reference set (Supplementary data online, *Methods S1*).^25^ The GDPPR dataset includes patients who are currently registered and deceased patients with a date of death on or after 1 November 2019. For patients in the GDPPR dataset, the available records at the time of data extraction included diagnoses up to 31 July 2024. GP records transfer between practices when a patient moves, and so should capture an individual’s entire medical history. We also used the Hospital Episode Statistics Admitted Patient Care (HES APC) dataset, which included inpatient admissions and day cases from 1 April 1997 to 31 July 2024. The HES APC dataset includes the primary diagnosis (main reason for hospitalisation) up to 19 secondary diagnoses and up to 24 procedures. Data from the Civil Registrations of Death, which includes all deaths registered in England, was available from 1 November 2019 to 31 July 2024 and we extracted date of death and underlying cause of death. Finally, we also used data from COVID-19 Hospitalisations in England Surveillance System (CHESS), Second Generation Surveillance System (SGSS) and Secondary Uses Service (SUS), all of which can be used to identify people with COVID-19 infection.

### Study design and study population

We conducted a retrospective cohort study, including all adults (≥ 18 years of age) with a final diagnosis of NSTEMI or STEMI recorded in MINAP between 1 November 2019 to 28 February 2023. We did not include events prior to 1 November 2019, since GDPPR data was unavailable for people who died before this date. We excluded patients without a linked GDPPR record and those who failed quality assurance checks. For each individual we included the first record of an MI within the study period.

### Mental disorder

We ascertained history of schizophrenia, bipolar disorder and depression (of any severity) from the GDPPR and HES APC datasets, based on diagnoses at any time prior to each individual’s MI admission date. We categorised people into mutually exclusive groups using a severity hierarchy, with schizophrenia considered the most severe, followed by bipolar disorder and depression. For the HES APC dataset, we used ICD-10 codes, as per previous research^4,26^, to identify schizophrenia (F20, F25), bipolar disorder (F30, F31) and depression (F32, F33) from the primary and secondary diagnosis fields. For the GDPPR dataset, we identified each disorder using SNOMED codes (Supplementary data online, *Methods S1*). The comparison group included people without a diagnosis of any of these mental disorders prior to their MI admission date.

### Outcomes

Our primary outcome was 30-day mortality, measured from the MI admission date. We also evaluated: one-year mortality; one-year mortality amongst patients who survived 30 days (in order to evaluate whether any differences in one-year mortality were due to differences within and/or beyond the first 30 days); time to mortality within the first year; and time to cardiovascular disease (CVD) mortality within the first year. We defined CVD mortality based on deaths where the underlying cause was IHD (ICD-10 codes I20-I25) or cerebrovascular disease (I60-I69, G45). Finally, we evaluated one-year mortality amongst patients discharged home, adjusting for receipt of guideline-indicated hospital care indicators recommended for all such patients.

### Covariates

We ascertained age, sex, deprivation (based on quintiles of the Index of Multiple Deprivation,^27^ a neighbourhood-level measure of deprivation) and ethnicity from the CVD-COVID-UK/COVID-IMPACT Consortium key patient characteristics table.^28^ We defined MI admission timing variables: daytime or overnight; weekday or weekend and 4-month period during which the admission occurred; and identified the hospital where the patient was admitted (Supplementary data online, *Methods S2*).

We identified history of MI and comorbid angina, cerebrovascular disease, chronic renal failure, diabetes and heart failure from the MINAP dataset and from diagnoses recorded in GDPPR or HES APC prior to the date of MI admission, based on SNOMED and ICD-10 code lists^4,23,29–33^ (Supplementary data online, *Methods S2*). We identified history of PCI from the MINAP dataset and from procedures recorded in HES APC using an existing OPCS-4 code list^4^ (Supplementary data online, *Methods S2*). We ascertained MI presentation features including cardiac arrest, cardiogenic shock, ST deviation, creatinine within the first 24 hours after admission (μmol/L), heart rate on admission and the first systolic blood pressure (SBP) (mmHg) measurement after admission from the MINAP dataset. We identified patients who had COVID-19 within 14 days either side of their MI (Supplementary data online, *Methods S2*). We also defined guideline-informed care standards relevant to patients discharged home, specifically receipt of angiography and admission to cardiac ward for NSTEMI, receipt of primary PCI and call-to-balloon (CTB) time (i.e. time from call for help to receipt of primary PCI) within target time for STEMI, and indicated secondary prevention medication prescribed at discharge and referral for cardiac rehabilitation for both types of MI (Supplementary data online, *Methods S2*).

### Statistical analyses

We analysed NSTEMI and STEMI separately, using mixed effects logistic regression for the binary outcomes and mixed effects Cox proportional hazards models for the time-to-event outcomes. For time to CVD mortality within the first year, we accounted for the competing risk of death from other causes by censoring people who died from other causes at their date of death. We obtained odds ratios (ORs) (for binary outcomes) and hazard ratios (HRs) (for time-to-event outcomes), comparing patients with each of schizophrenia, bipolar disorder or depression to those without any of these disorders.

For each of NSTEMI and STEMI, we serially adjusted for groups of potential confounding factors: model 1 adjusted for age and sex, with random effects for the intercept for each hospital; model 2 additionally adjusted for ethnicity and deprivation; model 3 additionally adjusted for timing variables; model 4 additionally adjusted for clinical history (history of MI, history of PCI and comorbidities); and model 5 additionally adjusted for MI presentation features (cardiac arrest, cardiogenic shock, creatinine, heart rate, SBP and COVID-19 status for both types of MI, and additionally ST deviation for NSTEMI only). For one-year mortality amongst patients discharged home, we additionally sequentially adjusted for guideline-informed care standards. Age, creatinine, heart rate and SBP were included in the models as continuous variables. We scaled age, SBP and heart rate by subtracting their respective means and dividing by their standard deviations.

Creatinine was right skewed and hence was log transformed prior to scaling to improve convergence of the models. For all continuous variables, we included both linear and quadratic terms in the models because exploratory plots suggested that quadratic functions were sufficient for describing the relationships between these variables and the outcomes. For the mixed effects Cox proportional hazards models, we used log cumulative hazard plots to check the proportional hazards assumption. We investigated whether any associations between mental disorders and mortality were affected by the COVID-19 pandemic. Given that our study data were only available from November 2019, we evaluated the effect of the pandemic by comparing the associations in the pre-pandemic period (November 2019 – February 2020) to the associations in each of the following 4-month periods. We categorised the time period into four-month intervals because we wanted to capture fluctuations in mortality throughout our study period without making prior assumptions that specific periods (for example, periods with high COVID-19 mortality) would have higher or lower mortality rates. In addition to periods with high COVID-19 mortality, mortality throughout our study period may have been affected by lockdowns and periods of increased demand on ambulance services and hospitals. For the analysis of the impact of the COVID-19 pandemic, we used a binary mental disorder variable (any of schizophrenia, bipolar disorder or depression versus none of these disorders) since there were insufficient numbers to investigate the interaction for individual mental disorders. We refitted the final model for each outcome using the binary mental disorder variable instead of the original categorical variable and fitted a further model including the interaction between the binary mental disorder variable and 4-month time period. We examined whether the interaction term improved model fit using a likelihood ratio test.

The primary analyses excluded patients with missing data in the outcome or any of the covariates. Missing data was minimal amongst the sociodemographic, timing and clinical history covariates, but higher amongst the MI presentation covariates. Hence, we also conducted a post hoc sensitivity analysis that omitted the MI presentation covariates from the sequence of models and excluded only patients with missing data in this narrower set of covariates.

In accordance with statistical disclosure control rules, counts are rounded to the nearest 5 and counts less than 10 are suppressed. We used R (version 4.1.3 and above)^34^ to conduct the statistical analysis, the package lme4^35^ to fit mixed effects logistic regression models and the package coxme^36^ to fit mixed effects Cox proportional hazards models. All R code is available in our GitHub repository (https://github.com/BHFDSC/CCU046_02).

## Results

### Descriptive findings

During the study period, 131 075 patients with an NSTEMI and 79 045 with a STEMI were eligible for inclusion in our study (Figure 1, Supplementary data online, *Figure S1*). Among patients with an NSTEMI, 875 (0.7%), 750 (0.6%) and 34 610 (26.4%) had a prior record of schizophrenia, bipolar disorder and depression, respectively, with prevalence similar among those with a STEMI.

**Figure 1:**
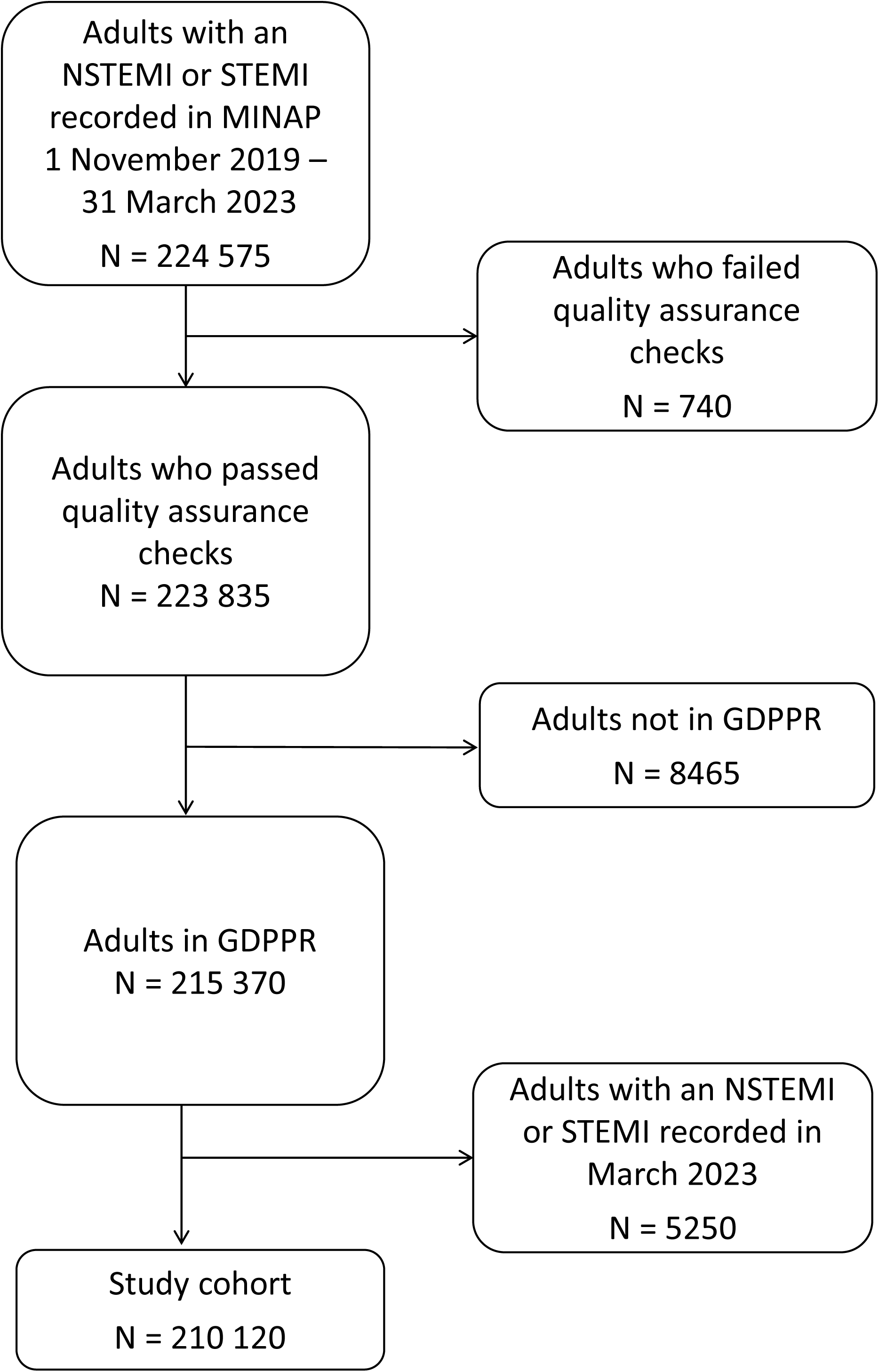
Flow diagram for the study cohort.

For both NSTEMI and STEMI, people with versus without a mental disorder diagnosis were: younger at MI onset; more likely to be women; and more likely to live in the most deprived areas. Minority ethnic groups were over-represented among patients with schizophrenia, and under-represented among patients with bipolar disorder or depression, relative to the comparison group. People with a mental disorder diagnosis were more likely to be admitted overnight and generally had a higher prevalence of cardiovascular disease, cerebrovascular disease and diabetes. MI presentation features were broadly similar across groups (Tables 1 and 2).

**Table 1.** Baseline characteristics of patients with a MINAP record of an NSTEMI, by mental disorder status^a^.

| Characteristic | Schizophrenia<br>(N = 875) | Bipolar disorder<br>(N = 750) | Depression<br>(N = 34 610) | None of these disorders<br>(N =94 840) |
| --- | --- | --- | --- | --- |
| **Sociodemographics** |  |  |  |  |
| Age at MI (years), mean (SD) | 64.7 (12.5) | 65.3 (12.9) | 67.5 (13.0) | 70.6 (13.3) |
| Female (%) | 295 (33.7) | 320 (42.7) | 15 170 (43.8) | 27 840 (29.4) |
| IMD decile (%) |  |  |  |  |
| 1 (most deprived) | 195 (22.3) | 120 (16.0) | 5210 (15.1) | 9710 (10.2) |
| 2 | 155 (17.7) | 105 (14.0) | 4155 (12.0) | 9150 (9.6) |
| 3 | 115 (13.1) | 75 (10.0) | 3830 (11.1) | 9290 (9.8) |
| 4 | 105 (12.0) | 105 (14.0) | 3655 (10.6) | 9570 (10.1) |
| 5 | 70 (8.0) | 65 (8.7) | 3470 (10.0) | 9620 (10.1) |
| 6 | 65 (7.4) | 70 (9.3) | 3275 (9.5) | 9805 (10.3) |
| 7 | 45 (5.1) | 75 (10.0) | 3100 (9.0) | 9535 (10.1) |
| 8 | 55 (6.3) | 45 (6.0) | 2870 (8.3) | 9425 (9.9) |
| 9 | 35 (4.0) | 55 (7.3) | 2645 (7.6) | 9210 (9.7) |
| 10 (least deprived) | 30 (3.4) | 40 (5.3) | 2380 (6.9) | 9410 (9.9) |
| Missing | <10 (<1.1) | <10 (<1.3) | 25 (0.1) | 110 (0.1) |
| Ethnicity |  |  |  |  |
| Black | 30 (3.4) | 10 (1.3) | 365 (1.1) | 1700 (1.8) |
| South Asian | 125 (14.3) | 50 (6.7) | 1785 (5.2) | 7990 (8.4) |
| White | 680 (77.7) | 675 (90.0) | 31 535 (91.1) | 81 330 (85.8) |
| Mixed | <10 (<1.1) | <10 (<1.3) | 240 (0.7) | 685 (0.7) |
| Other ethnic groups | 30 (3.4) | 10 (1.3) | 675 (2.0) | 2970 (3.1) |
| Missing | <10 (<1.1) | <10 (<1.3) | 10 (<0.1) | 170 (0.2) |
| **Location and timing** |  |  |  |  |
| NHS trust |  |  |  |  |
| Available | 855 (97.7) | 725 (96.7) | 33 660 (97.3) | 92 260 (97.3) |
| Missing | 20 (2.3) | 25 (3.3) | 955 (2.8) | 2580 (2.7) |
| Admission day |  |  |  |  |
| Week day | 670 (76.6) | 565 (75.3) | 26 030 (75.2) | 71 370 (75.3) |
| Weekend | 205 (23.4) | 190 (25.3) | 8580 (24.8) | 23 470 (24.7) |
| Admission timing |  |  |  |  |
| Daytime | 485 (55.4) | 425 (56.7) | 20 815 (60.1) | 58 720 (61.9) |
| Overnight | 390 (44.6) | 330 (44.0) | 13 795 (39.9) | 36 120 (38.1) |
| **Clinical history** |  |  |  |  |
| Previous MI (%) | 235 (26.9) | 215 (28.7) | 9220 (26.6) | 21 755 (22.9) |
| Previous PCI (%) | 200 (22.9) | 210 (28.0) | 10 525 (30.4) | 27 310 (28.8) |
| Angina (%) | 420 (48.0) | 390 (52.0) | 17 200 (49.7) | 42 710 (45.0) |
| Cerebrovascular disease (%) | 225 (25.7) | 195 (26.0) | 7105 (20.5) | 14 980 (15.8) |
| Chronic renal failure (%) | 100 (11.4) | 85 (11.3) | 3455 (10.0) | 9750 (10.3) |
| Diabetes (%) | 430 (49.1) | 320 (42.7) | 13 780 (39.8) | 33 475 (35.3) |
| Heart failure (%) | 300 (34.3) | 215 (28.7) | 10 030 (29.0) | 25 700 (27.1) |
| **MI presentation features** |  |  |  |  |
| ST deviation (%) | 170 (19.4) | 130 (17.3) | 6585 (19.0) | 19 320 (20.4) |
| Missing (%) | 30 (3.4) | 15 (2.0) | 1140 (3.3) | 2810 (3.0) |
| Cardiogenic shock (%) | <10 (<1.1) | <10 (<1.3) | 125 (0.4) | 410 (0.4) |
| Missing (%) | 115 (13.1) | 75 (10.0) | 3820 (11.0) | 10 310 (10.9) |
| Cardiac arrest (%) | 35 (4.0) | 15 (2.0) | 755 (2.2) | 2445 (2.6) |
| Missing (%) | 25 (2.9) | 10 (1.3) | 660 (1.9) | 1880 (2.0) |
| Creatinine (μmol/L) |  |  |  |  |
| Median [IQR] | 83.0 [68.0, 104.0] | 83.0 [68.0, 104.0] | 82.0 [68.0, 101.0] | 85.0 [72.0, 105.0] |
| Missing (%) | 35 (4.0) | 20 (2.7) | 970 (2.8) | 2475 (2.6) |
| SBP (mmHg) |  |  |  |  |
| Mean (SD) | 138.2 (27.5) | 140.0 (28.2) | 143.3 (27.3) | 145.1 (27.4) |
| Missing (%) | 35 (4.0) | 15 (2.0) | 795 (2.3) | 2340 (2.5) |
| Heart rate (bpm) |  |  |  |  |
| Mean (SD) | 85.8 (20.4) | 82.8 (19.8) | 80.7 (19.0) | 79.8 (19.2) |
| Missing (%) | 30 (3.4) | 15 (2.0) | 830 (2.4) | 2420 (2.6) |
| COVID-19 positive (%) | 50 (5.7) | 45 (6.0) | 1715 (5.0) | 4995 (5.3) |

<sup>a</sup>As per the statistical disclosure control rules, counts are rounded to the nearest 5 and counts less than 10 are suppressed bpm, beats per minute; IMD, index of multiple deprivation; IQR, interquartile range; MI, myocardial infarction; PCI, percutaneous coronary intervention; SBP, systolic blood pressure; SD, standard deviation.

**Table 2.** Baseline characteristics of patients with a MINAP record of a STEMI, by mental disorder status^a^.

|  | Schizophrenia<br>(N = 490) | Bipolar disorder<br>(N = 405) | Depression<br>(N = 19 230) | None of these disorders<br>(N = 58 920) |
| --- | --- | --- | --- | --- |
| **Sociodemographics** |  |  |  |  |
| Age at MI (years), mean (SD) | 60.3 (13.1) | 61.6 (13.0) | 63.6 (12.8) | 66.5 (13.3) |
| Female (%) | 130 (26.5) | 170 (42.0) | 7200 (37.4) | 13 995 (23.8) |
| IMD decile (%) |  |  |  |  |
| 1 (most deprived) | 95 (19.4) | 65 (16.0) | 3040 (15.8) | 6280 (10.7) |
| 2 | 85 (17.3) | 45 (11.1) | 2450 (12.7) | 5930 (10.1) |
| 3 | 70 (14.3) | 50 (12.3) | 2180 (11.3) | 5800 (9.8) |
| 4 | 60 (12.2) | 45 (11.1) | 2055 (10.7) | 5995 (10.2) |
| 5 | 45 (9.2) | 40 (9.9) | 1860 (9.7) | 5980 (10.1) |
| 6 | 30 (6.1) | 20 (4.9) | 1750 (9.1) | 5970 (10.1) |
| 7 | 30 (6.1) | 40 (9.9) | 1590 (8.3) | 5855 (9.9) |
| 8 | 30 (6.1) | 50 (12.3) | 1560 (8.1) | 5925 (10.1) |
| 9 | 25 (5.1) | 25 (6.2) | 1495 (7.8) | 5745 (9.8) |
| 10 (least deprived) | 15 (3.1) | 20 (4.9) | 1240 (6.4) | 5405 (9.2) |
| Missing | <10 (<2.0) | <10 (<2.5) | 10 (0.1) | 45 (0.1) |
| Ethnicity |  |  |  |  |
| Black | 15 (3.1) | <10 (<2.5) | 180 (0.9) | 905 (1.5) |
| South Asian | 50 (10.2) | 15 (3.7) | 825 (4.3) | 4950 (8.4) |
| White | 390 (79.6) | 375 (92.6) | 17 710 (92.1) | 50 640 (85.9) |
| Mixed | 10 (2.0) | <10 (<2.5) | 125 (0.7) | 425 (0.7) |
| Other ethnic groups | 20 (4.1) | <10 (<2.5) | 360 (1.9) | 1750 (3.0) |
| Missing | <10 (<2.0) | <10 (<2.5) | 30 (0.2) | 250 (0.4) |
| **Location and timing** |  |  |  |  |
| NHS trust (%) |  |  |  |  |
| Available | 485 (99.0) | 390 (96.3) | 18 945 (98.5) | 58 045 (98.5) |
| Missing | <10 (<2.0) | 15 (3.7) | 285 (1.5) | 875 (1.5) |
| Admission day |  |  |  |  |
| Week day | 355 (72.4) | 305 (75.3) | 13 810 (71.8) | 42 220 (71.7) |
| Weekend | 135 (27.6) | 100 (24.7) | 5420 (28.2) | 16 700 (28.3) |
| Admission timing |  |  |  |  |
| Daytime | 265 (54.1) | 225 (55.6) | 11 550 (60.1) | 36 845 (62.5) |
| Overnight | 225 (45.9) | 180 (44.4) | 7675 (39.9) | 22 075 (37.5) |
| **Clinical history** |  |  |  |  |
| Previous MI (%) | 85 (17.3) | 65 (16.0) | 3035 (15.8) | 6905 (11.7) |
| Previous PCI (%) | 315 (64.3) | 285 (70.4) | 14 715 (76.5) | 44 735 (75.9) |
| Angina (%) | 130 (26.5) | 120 (29.6) | 5885 (30.6) | 14 830 (25.2) |
| Cerebrovascular disease (%) | 80 (16.3) | 80 (19.8) | 2565 (13.3) | 5815 (9.9) |
| Chronic renal failure (%) | 20 (4.1) | 25 (6.2) | 815 (4.2) | 2340 (4.0) |
| Diabetes (%) | 190 (38.8) | 145 (35.8) | 6075 (31.6) | 15 530 (26.4) |
| Heart failure (%) | 180 (36.7) | 115 (28.4) | 6265 (32.6) | 18 655 (31.7) |
| **MI presentation features** |  |  |  |  |
| Cardiogenic shock (%) | 25 (5.1) | 15 (3.7) | 580 (3.0) | 2130 (3.6) |
| Missing (%) | 75 (15.3) | 40 (9.9) | 2700 (14.0) | 8150 (13.8) |
| Cardiac arrest (%) | 60 (12.2) | 50 (12.3) | 2290 (11.9) | 7630 (12.9) |
| Missing (%) | 15 (3.1) | 10 (2.5) | 355 (1.8) | 1330 (2.3) |
| Creatinine (micromole/L) |  |  |  |  |
| Median [IQR] | 80.0 [67.0, 99.0] | 78.0 [64.2, 99.0] | 78.0 [65.0, 94.0] | 81.0 [69.0, 98.0] |
| Missing (%) | 30 (6.1) | 20 (4.9) | 1230 (6.4) | 3310 (5.6) |
| SBP (mmHg) |  |  |  |  |
| Mean (SD) | 129.3 (27.2) | 128.8 (26.3) | 132.7 (27.7) | 133.0 (27.7) |
| Missing (%) | 30 (6.1) | 15 (3.7) | 755 (3.9) | 2410 (4.1) |
| Heart rate (bpm) |  |  |  |  |
| Mean (SD) | 85.2 (20.8) | 82.0 (19.5) | 80.1 (19.3) | 79.0 (19.3) |
| Missing | 25 (5.1) | 10 (2.5) | 680 (3.5) | 2210 (3.8) |
| COVID-19 positive (%) | 25 (5.1) | 15 (3.7) | 835 (4.3) | 2595 (4.4) |

<sup>a</sup>As per the statistical disclosure control rules, counts are rounded to the nearest 5 and counts less than 10 are suppressed bpm, beats per minute; IMD, index of multiple deprivation; IQR, interquartile range; MI, myocardial infarction; PCI, percutaneous coronary intervention; SBP, systolic blood pressure; SD, standard deviation.

### Mental disorder and 30-day and one-year mortality

Among patients with NSTEMI, 6600 (5.0%) died within 30 days, with the crude percentage highest among those with schizophrenia. Among those with STEMI, 7440 (9.4%) died within 30 days, with the crude percentages highest amongst those with schizophrenia or bipolar disorder. For both types of MI, the proportion of patients who died within one year was greater among those with schizophrenia and bipolar disorder than in the comparison group (Table 3).

**Table 3.** Mortality and receipt of care among people with each of NSTEMI and STEMI, by mental disorder status^a^.

| Mortality | Schizophrenia | Bipolar disorder | Depression | None of these disorders |
| --- | --- | --- | --- | --- |
| **NSTEMI** |  |  |  |  |
| All patients (N) | 875 | 750 | 34 610 | 94 840 |
| 30-day all-cause mortality (%) | 60 (6.9) | 35 (4.7) | 1625 (4.7) | 4875 (5.1) |
| 1-year all-cause mortality (%) | 165 (18.9) | 115 (15.3) | 4540 (13.1) | 12 915 (13.6) |
| 1-year CVD mortality (%) | 85 (9.7) | 65 (8.7) | 2320 (6.7) | 6305 (6.6) |
| Patients who survived 30 days (N) | 815 | 715 | 32 990 | 89 960 |
| 1-year all-cause mortality (%) | 100 (12.3) | 80 (11.2) | 2920 (8.9) | 8035 (8.9) |
| Patients discharged home (N) | 660 | 535 | 26 210 | 71 420 |
| 1-year all-cause mortality (%) | 100 (15.2) | 70 (13.1) | 2805 (10.7) | 7700 (10.8) |
| Angiogram (%) |  |  |  |  |
| Received within 72 hours | 185 (28.0) | 200 (37.4) | 10 590 (40.4) | 29 835 (41.8) |
| Received out with 72 hours | 155 (23.5) | 135 (25.2) | 6805 (26.0) | 18 660 (26.1) |
| Received, timing unknown | 30 (4.5) | 25 (4.7) | 1485 (5.7) | 3925 (5.5) |
| Eligible, but no receipt | 155 (23.5) | 85 (15.9) | 3240 (12.4) | 8495 (11.9) |
| Not eligible | 110 (16.7) | 65 (12.1) | 3275 (12.5) | 8465 (11.9) |
| Missing | 30 (4.5) | 20 (3.7) | 815 (3.1) | 2045 (2.9) |
| Referred for cardiac rehabilitation (%) |  |  |  |  |
| Yes | 420 (63.6) | 385 (72.0) | 20 265 (77.3) | 55 435 (77.6) |
| No | 80 (12.1) | 45 (8.4) | 2125 (8.1) | 5845 (8.2) |
| Not indicated | 85 (12.9) | 50 (9.3) | 1910 (7.3) | 5075 (7.1) |
| Patient declined | 20 (3.0) | 10 (1.9) | 380 (1.4) | 820 (1.1) |
| Missing | 55 (8.3) | 40 (7.5) | 1535 (5.9) | 4250 (6.0) |
| Discharged on secondary prevention (%) |  |  |  |  |
| Yes | 535 (81.1) | 440 (82.2) | 21 830 (83.3) | 60 200 (84.3) |
| No | 100 (15.2) | 80 (15.0) | 3630 (13.8) | 9410 (13.2) |
| Missing | 25 (3.8) | 15 (2.8) | 750 (2.9) | 1810 (2.5) |
| Admission to cardiac ward (%) |  |  |  |  |
| Yes | 260 (39.4) | 210 (39.3) | 11 110 (42.4) | 30 810 (43.1) |
| No | 380 (57.6) | 305 (57.0) | 14 500 (55.3) | 38 925 (54.5) |
| Missing | 20 (3.0) | 15 (2.8) | 600 (2.3) | 1685 (2.4) |
| **STEMI** |  |  |  |  |
| All patients (N) | 490 | 405 | 19 230 | 58 920 |
| 30-day all-cause mortality (%) | 60 (12.2) | 50 (12.3) | 1705 (8.9) | 5625 (9.5) |
| 1-year all-cause mortality (%) | 90 (18.4) | 70 (17.3) | 2570 (13.4) | 8190 (13.9) |
| 1-year CVD mortality (%) | 65 (13.3) | 50 (12.3) | 1820 (9.5) | 5970 (10.1) |
| Patients who survived 30 days (N) | 430 | 350 | 17520 | 53 300 |
| 1-year all-cause mortality (%) | 35 (8.1) | 15 (4.3) | 860 (4.9) | 2565 (4.8) |
| Patients discharged home (N) | 380 | 330 | 16 300 | 49 795 |
| 1-year all-cause mortality (%) | 30 (7.9) | 20 (6.1) | 930 (5.7) | 2820 (5.7) |
| Call-to-balloon |  |  |  |  |
| <120 minutes | 105 (27.6) | 60 (18.2) | 4165 (25.6) | 13 370 (26.9) |
| 120-150 minutes | 55 (14.5) | 65 (19.7) | 3320 (20.4) | 9970 (20.0) |
| ≥150 minutes | 105 (27.6) | 115 (34.8) | 4955 (30.4) | 14 530 (29.2) |
| Received PCI, but CTB time unknown | 35 (9.2) | 30 (9.1) | 1295 (7.9) | 4240 (8.5) |
| Did not receive PCI | 80 (21.1) | 55 (16.7) | 2495 (15.3) | 7465 (15.0) |
| Missing | <10 (<2.6) | <10 (<3.0) | 75 (0.5) | 225 (0.5) |
| Referred for cardiac rehabilitation (%) |  |  |  |  |
| Yes | 320 (84.2) | 290 (87.9) | 14 805 (90.8) | 44 980 (90.3) |
| No | 25 (6.6) | 15 (4.5) | 490 (3.0) | 1730 (3.5) |
| Not indicated | 20 (5.3) | <10 (<3.0) | 335 (2.1) | 1075 (2.2) |
| Patient declined | 10 (2.6) | <10 (<3.0) | 100 (0.6) | 265 (0.5) |
| Missing | 10 (2.6) | 15 (4.5) | 575 (3.5) | 1745 (3.5) |
| Discharged on secondary prevention (%) |  |  |  |  |
| Yes | 340 (89.5) | 305 (92.4) | 15 085 (92.5) | 46 205 (92.8) |
| No | 25 (6.6) | 15 (4.5) | 805 (4.9) | 2525 (5.1) |
| Missing | 15 (3.9) | <10 (<3.0) | 410 (2.5) | 1065 (2.1) |

<sup>a</sup>In accordance with NHSE's SDE statistical disclosure control rules, counts are rounded to the nearest 5 and counts less than 10 are suppressed

After adjusting for sociodemographic and timing covariates, clinical history and MI presentation features, relative to the comparison group, odds of 30-day mortality following NSTEMI were 73% higher in people with schizophrenia (OR 1.73, 1.20–2.49) and 17% higher in those with depression (1.17, 1.08–1.26). For bipolar disorder there was not a statistically significant difference in the odds of 30-day mortality (OR 1.17, 0.75-1.84). (Figure 2; Supplementary data online *Figure S2*).

**Figure 2:**
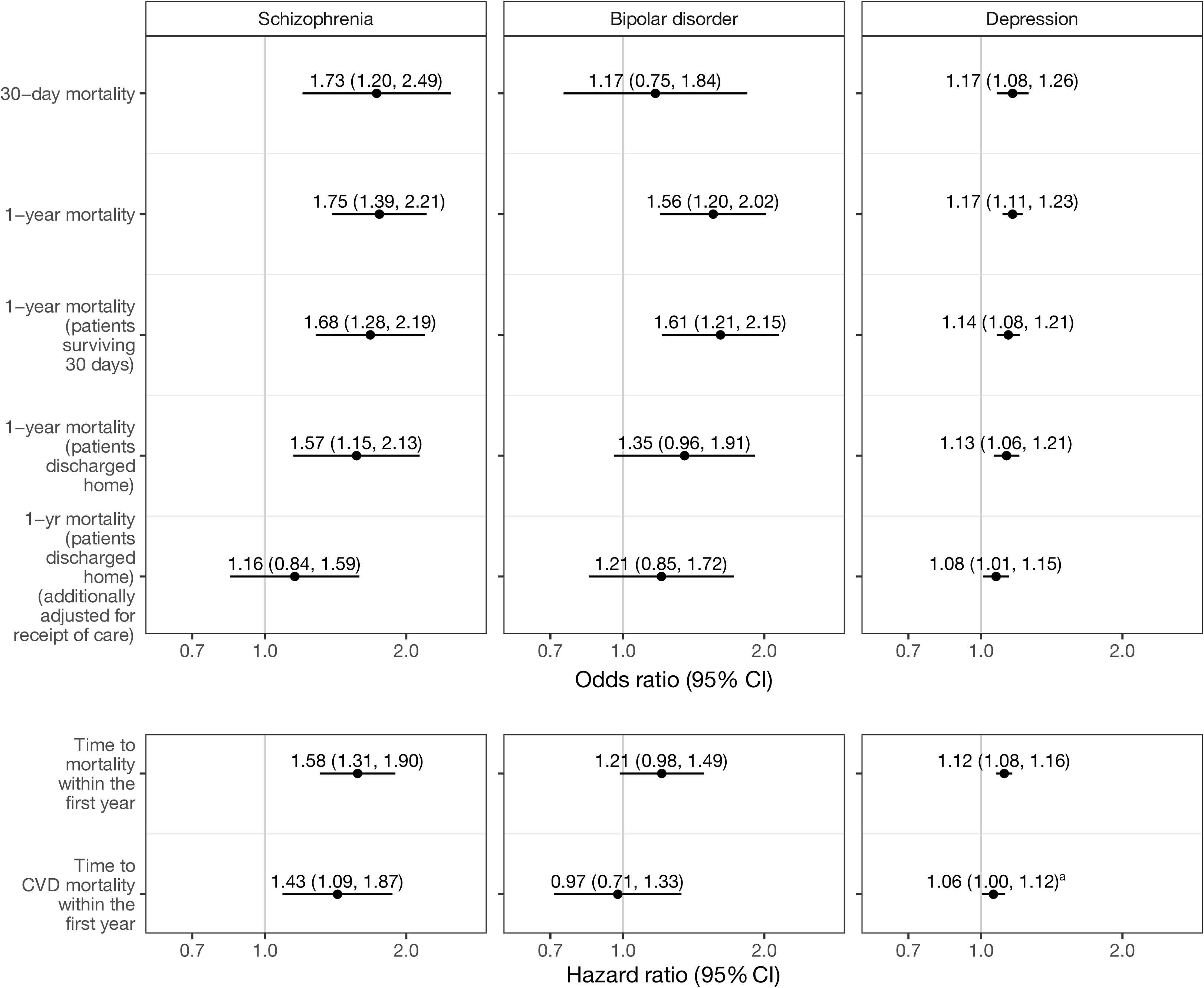
Odds ratios and hazard ratios for mortality following NSTEMI in patients with schizophrenia, bipolar disorder or depression versus those without any of these disorders. Ratios are adjusted for sociodemographic characteristics, hospital, MI timing, comorbidities and MI presentation features (model 5). ^a^1.062 (1.004, 1.123)

For STEMI, after adjustment for all covariates, 30-day mortality was higher for people with schizophrenia (OR 1.61, 1.09–2.37), bipolar disorder (OR 1.68, 1.08–2.59) and depression (OR 1.10, 1.01–1.20) (Figure 3; Supplementary data online, *Figure S3*).

**Figure 3:**
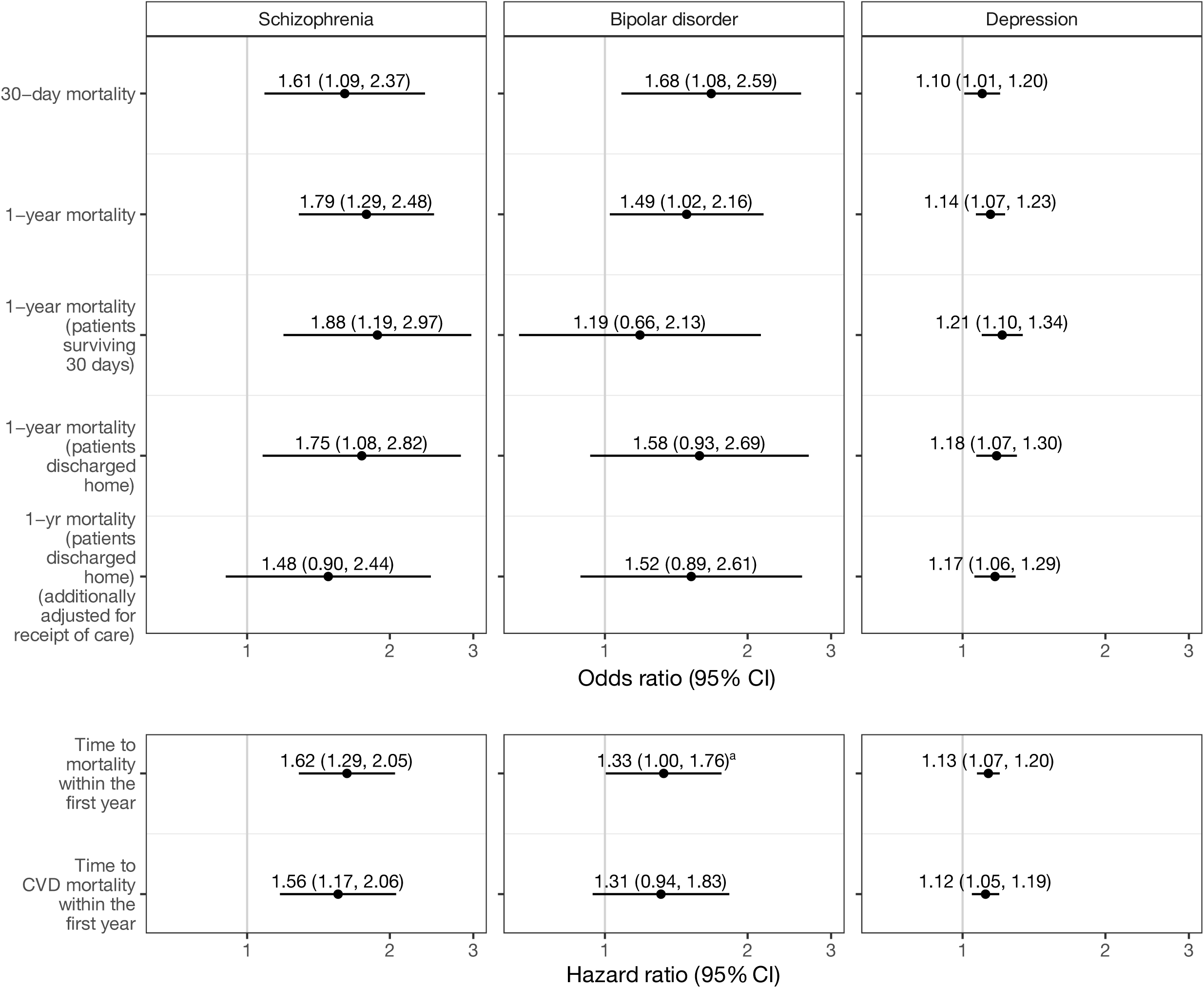
Odds ratios and hazard ratios for mortality following NSTEMI in patients with schizophrenia, bipolar disorder or depression versus those without any of these disorders. Ratios are adjusted for sociodemographic characteristics, hospital, MI timing, comorbidities and MI presentation features (model 5). ^a^1.331 (1.005, 1.762)

For both types of MI, each mental disorder was associated with excess odds of one-year mortality after adjustment for all covariates. Among patients with an NSTEMI, schizophrenia, bipolar disorder and depression were associated with increases of 75% (OR 1.75, 1.39–2.21), 56% (OR 1.56, 1.20–2.02) and 17% (OR 1.17, 1.11–1.23), respectively (Figure 2). ORs for the association between mental disorder and one-year mortality following STEMI were similar to those for NSTEMI (Figure 3; Supplementary data online, *Figures S2 and S3*).

Disparities in 1-year mortality were not entirely attributable to disparities within the first 30 days. Amongst patients with NSTEMI who survived 30 days, fully adjusted models still indicated higher odds of 1-year mortality for people with each of the disorders, relative to the comparison group (Figure 2; Supplementary data online, *Figure S2*). Amongst STEMI patients who survived 30 days, the fully adjusted models indicated higher odds of 1-year mortality for people with schizophrenia or depression, but not for bipolar disorder, where confidence intervals were extremely wide (Figure 3; Supplementary data online, *Figure S3*).

For time to mortality within 1 year, based on the fully adjusted models and relative to the comparison group, survival time was lower in NSTEMI patients with schizophrenia or depression, and in STEMI patients with any of schizophrenia, bipolar disorder or depression (Figures 2 and 3; Supplementary data online, *Figures S2 and S3*).

For both types of MI, based on the fully adjusted models, HRs for cardiovascular death within one year were statistically significantly higher in people with schizophrenia and (to a much lesser extent) depression, relative to the comparison group. For bipolar disorder there was no clear evidence of a difference in time to cardiovascular death relative to the comparison group (Figures 2 and 3; Supplementary data online, *Figures S2 and S3*).

### Exploration of effect of receipt of care on one-year mortality

Among patients with NSTEMI who were discharged home, those with schizophrenia, bipolar disorder or depression were less likely to receive guideline indicated care (Table 3). In models of one-year mortality, additional adjustment for receipt of care indicators attenuated effect estimates, particularly for schizophrenia (Figure 2). The OR for the association between schizophrenia and one-year mortality attenuated from 1.57 (95% CI 1.15–2.13) to 1.16 (95% CI 0.84–1.59).

Among STEMI patients who were discharged home, those with schizophrenia, bipolar disorder or depression were less likely to receive PCI, and those with schizophrenia were less likely to be referred for cardiac rehabilitation and be discharged on all indicated secondary prevention medication. After adjusting one-year mortality for receipt of care, effect estimates for schizophrenia attenuated from OR 1.75 (95% CI 1.08–2.82) to OR 1.48 (95% CI 0.90–2.44). Estimates attenuated only slightly for depression and bipolar disorder (Figure 3).

Individual adjustment for care indicators revealed that most of the attenuation of effect estimates arose from adjusting for receipt of angiogram in NSTEMI, and adjusting for receipt of primary PCI and referral for cardiac rehabilitation in STEMI (Supplementary data online, *Figures S2 and S3*).

### Sensitivity analyses

Findings from models that included patients with complete information on sociodemographic, timing and clinical history variables (97% and 98% of all patients eligible for our NSTEMI and STEMI cohorts, respectively) but did not adjust for MI presentation features were similar to those obtained in the primary analysis (Supplementary data online, *Figures S4 and S5*).

### COVID-19 and mental disorder differences in outcomes

Within the study period, overall monthly admissions for both NSTEMI and STEMI were lowest at the start of the COVID-19 pandemic, in March and April 2020 (Supplementary data online, *Figure S6*). For NSTEMI, 30-day mortality was highest from March to April 2020, and from December 2020 to January 2021. For STEMI, 30-day mortality was highest in March 2020, December 2020 and from December 2022 to January 2023. Time trends were similar for patients with and without a mental disorder diagnosis (Supplementary data online, *Figure S7*). There was no evidence that the associations between mental disorder and mortality varied over our study period, with one exception; for NSTEMI, the association between mental disorder and time to CVD mortality within the first year varied by admission period (Supplementary data online, *Table S1*). There was a statistically significant difference between the association in the pre-pandemic period (November 2019 – February 2020), and the association in one other period (November 2020 – February 2021). In this period only, people with schizophrenia, bipolar disorder or depression had a longer time to CVD mortality than the comparison group (Supplementary data online, *Table S2*). Findings from the sensitivity analysis were similar, with the association between mortality and mental disorder only varying by period for time to CVD mortality amongst people with NSTEMI (Supplementary data online, *Tables S3 and S4*).

## Discussion

We found that each of schizophrenia, bipolar disorder and depression were associated with higher one-year mortality following both an NSTEMI and STEMI, even after accounting for key confounders. The excess mortality was generally largest for schizophrenia, but was also evident for bipolar disorder, and to a lesser degree, depression. Additionally, 30-day mortality following NSTEMI was higher amongst patients with schizophrenia or depression and amongst STEMI patients with schizophrenia, bipolar disorder or depression, compared to those without these mental disorders.

Exploratory analyses revealed that differences in receipt of care may partly explain the excess one-year mortality among those with schizophrenia for both NSTEMI and STEMI. Within our study period, 30-day mortality following NSTEMI or STEMI generally peaked in line with the periods of highest mortality from COVID-19. Trends over time were similar for people with and without a mental disorder, hence disparities in MI mortality generally persisted, but did not widen, during and following the COVID-19 pandemic.

### Disparities by mental disorder

The 30-day and one-year mortality effect estimates generally align with previous findings on mental disorders and MI mortality risk from Scotland^4^ and other non-UK universal healthcare settings.^5,6,8,9,12^ Our study further adds to existing understanding by demonstrating similar patterns of excess mortality for both NSTEMI and STEMI. The generally higher excess risk of cardiovascular death in people with a mental disorder in our study aligns with reports of elevated risk of cardiovascular events post-MI in people with mental disorders.^4,17^ The finding that disparities are generally largest for schizophrenia is also consistent with previous studies reporting on multiple mental disorders within the same study population.^4,11,12^ Our findings make an important contribution to the neglected study of bipolar disorder and depression^4,11,12^ with respect to MI outcomes, highlighting disparities among people with mood disorders as well as psychotic disorders. The estimates for depression in the present study are smaller than those from studies where depression was ascertained from hospital admission records which are likely to predominantly capture more severe depression.^4,15^ A recent study supports possible variation in the association between depression and adverse MI outcomes by antidepressant medication prescribing, which potentially acts as a proxy for depression severity.^37^

Despite increasing evidence of MI mortality disparities by mental disorder status, there has been little advancement in understanding the underlying mechanisms. In our study, associations at both 30 days and one year persisted even after accounting for sociodemographic factors, timing of MI, clinical history and MI presentation features. Whilst the higher prevalence of comorbidities likely increases the risk of poorer outcome from MI in people with a mental disorder^38^, this may also indirectly operate through receipt of sub-optimal care.^39^ Studies have demonstrated that patients with comorbid diabetes, for example (particularly common in people with mental disorders) are less likely to receive optimal MI care.^40^ Our results suggest, that, for schizophrenia, differences in receipt of guideline-informed care standards for NSTEMI and STEMI translate into lower one-year survival even after taking account of differences in comorbidity prevalence. This is driven by lower access to reperfusion treatment and (for STEMI), lower referral for cardiac rehabilitation. These findings align with previous studies from Canada and Hong Kong reporting on schizophrenia^7^ and psychotic disorders^6^, with adjustment for receipt of reperfusion similarly attenuating one-year mortality estimates (although neither study stratified by MI type and focused on psychosis only).^7^ Additional factors, for which we did not have data or that were beyond scope of the present study, may also contribute to the observed differences in mortality. These include: presence of cardiac arrythmias, for which people with mental disorders are at a higher risk^41,42^; longer term prescription of cardiovascular secondary prevention medication, which is less likely in those with mental disorders^43,44^; and initiation (following referral) of cardiac rehabilitation, which may also be less likely in people with mental disorders since factors associated with initiating rehabilitation, such high socioeconomic status and fewer comorbidities, are less common in those with mental disorders.^45–47^ Finally, we did not examine secondary prevention medication adherence post-MI by mental disorder status, which is an understudied area. Whilst not specific to the post-MI period, one moderately sized study from the United States, found no evidence that people with schizophrenia were less likely to adhere to statins or angiotensin-converting enzyme inhibitor/angiotensin receptor blocker medications than people without psychiatric illness.^48^

### Impact of the COVID-19 pandemic

Mortality following NSTEMI or STEMI peaked in line with the periods of highest mortality from COVID-19 in the UK.^49^ To our knowledge, this is the first study to have examined the impact of the pandemic on disparities in post-MI mortality by mental disorder and, somewhat surprisingly, people with a mental disorder were not disproportionately affected.

During the early phase of the pandemic, the peak in 30-day mortality for both NSTEMI and STEMI in the total study population contrasts with an earlier study using MINAP data from January to May 2020, which reported higher 30-day mortality for NSTEMI but lower 30-day mortality for STEMI.^50^ However, various methodological differences, including differing time periods and inclusion criteria limit comparability and could account for these differing findings on 30-day mortality overall.

### Strengths and limitations

Strengths of this study include the use of a national and representative MI clinical care audit dataset with near-complete coverage.^24^ To our knowledge it is the first study to report on whether the COVID-19 pandemic affected existing mental disorder disparities in MI outcomes. Separate analysis of NSTEMI and STEMI and by mental disorder enabled a granular investigation of the mortality experience by type of MI and by individual mental disorder. In particular, this provides valuable insight into the MI mortality experience of people with bipolar disorder and depression, which has been little studied to date. Our study minimised information bias, through ascertainment of mental disorders from primary care records and linkage to national records for deaths. We also adjusted for various confounding factors, including sociodemographic factors and key comorbidities that influence MI survival. Finally, our exploratory assessment of the potential contribution of receipt of care standards across the acute care pathway makes a novel contribution to our understanding of the mechanisms underlying the mental disorder disparities in MI survival.

Our study has some limitations. Whilst the linked primary care data allowed better ascertainment of mental disorder, compared to previous studies based only on hospital records, we could not robustly determine severity of depression, since the most common depression codes recorded in primary care did not indicate severity. We had planned to focus our study on severe mental illness (schizophrenia, bipolar disorder and major depression), in order to evaluate mortality disparities in people with the most severe mental disorders, however due to the lack of specificity in primary care depression codes, we widened our definition of depression to include depression of any severity. Our findings for depression may therefore mask any heterogeneity of effect estimates by depression severity. Some misclassification of mental disorder status is likely, since we used a hierarchical approach to assign people with multiple diagnoses to their most severe disorder, and since people may not always seek help from their primary care providers for mental disorders, particularly depression. Evaluating post-MI morality disparities for other mental disorders such as generalised anxiety disorder and post-traumatic stress disorder was outside of the scope of this study, but is an important area for future research. Although the overall pattern of findings for bipolar disorder was generally similar to that for schizophrenia, effect estimates were less precise for bipolar disorder due to the smaller number with this disorder. Similarly, the smaller number of STEMI events led to reduced precision of effect estimates for mortality outcomes for bipolar disorder and schizophrenia. We performed complete case analysis rather than conducting multiple imputation given that 80% of cases were complete for the primary analysis and over 97% were complete in the sensitivity analyses where we excluded the MI presentation characteristics. Findings were similar for both the primary and the sensitivity analyses. Our effect estimates may be residually confounded by smoking, which is more common in people with mental disorders^51^ and associated with post-MI mortality^52^ and body mass index (BMI), which is higher in people with versus without mental disorders in the general population. Whilst the linked MINAP, hospital and primary care data available through the CVD-COVID-UK/COVID-IMPACT Consortium presented a significant opportunity to evaluate associations between mental disorder and mortality, our analysis of the impact of COVID-19 on these associations was limited since complete primary care data was only available from November 2019. With only four months of pre-pandemic data, we could not robustly account for seasonal fluctuations in mortality. Due to small numbers of people with schizophrenia and bipolar disorder, we combined the three mental disorders for the analyses of interaction with time period. However, given the much higher prevalence of depression than schizophrenia or bipolar disorder, this may have masked any variation in findings between disorders and we cannot conclusively rule out possible differences in mortality by time period for specific mental disorders.

### Implications

The attenuation of mortality estimates upon adjustment for receipt of care standards points to disparities in MI care as a modifiable contributory factor to mental disorder disparities in one-year survival. The residual excess risk also indicates that other factors play a role. Future research should focus on quantifying the contribution of acute care and other potential contributing factors, including post-discharge care, using appropriate mediation analysis approaches, to inform understanding of mechanisms and in turn development of tailored strategies. In-depth service evaluation, including interrogation of granular health data on MI symptom presentation, acute care pathways and in-hospital transitions is needed to identify the drivers of disparities in guideline-indicated acute care for MI. Qualitative studies of acute care for MI that include both the experiences of people with a mental disorder and an MI, and the experiences of the health care professionals who treat them, are also needed (with one such study from the authors of this paper underway). Given the established sociodemographic inequalities in cardiac care^54^, analysis of the intersection between mental disorders and factors such as sex, deprivation and ethnicity, is also warranted.

Meanwhile, health professionals involved in the delivery of acute cardiac care to people with mental disorders should be cognisant of the higher risk of MI in this group, the potential for sub-optimal treatment of MI, and the contribution of this to excess mortality in the short term. Better integration of care across clinical specialities during acute MI care, including emergency care, cardiology and liaison psychiatry would support higher quality care. Together with improved continuity of care across secondary and primary care, and improved medication review and cardiac rehabilitation referral and access processes, this could lead to reductions in the entrenched MI mortality disparities experienced by this vulnerable sub-population.

### Conclusions

We found that, in England, 30-day and one-year mortality are generally higher in people with each of schizophrenia, bipolar disorder and, to a lesser extent, depression and that these disparities did not worsen during and subsequent to the COVID-19 pandemic. Differences in receipt of care contributed to poorer survival at one year, reinforcing the urgent need for delivery of optimal acute cardiac care to people with a mental disorder.

## Author contributions

C.J. conceived the study. C.J., K.F., D.C., S.W.M, S.P and D.J.S. were awarded funding for the study. J.N. extracted the analysis dataset. K.F. conducted the statistical analysis. All authors contributed to the interpretation of the results. K.F. and C.J. wrote the first draft of the article. All authors commented on the draft and approved the final version.

## Acknowledgements

This work was carried out with the support of the BHF Data Science Centre led by HDR UK (BHF Grant no. SP/19/3/34678). This study made use of de-identified data held in NHS England’s Secure Data Environment service for England and made available via the BHF Data Science Centre’s CVD-COVID-UK/COVID-IMPACT consortium. The BHF DSC Health Data Science Team provided data curation resources and support. This work used data provided by patients and collected by the NHS as part of their care and support. We would also like to acknowledge all data providers who make health relevant data available for research.

## Data availability statement

The protocol, code lists and data curation and analysis code for this study are available on GitHub (https://github.com/BHFDSC/CCU046_02/).

The data used in this study are available in NHS England’s Secure Data Environment (SDE) service for England, but as restrictions apply they are not publicly available (https://digital.nhs.uk/services/secure-data-environment-service). The CVD-COVID-UK/COVID-IMPACT programme, led by the BHF Data Science Centre (https://bhfdatasciencecentre.org/), received approval to access data in NHS England’s SDE service for England from the Independent Group Advising on the Release of Data (IGARD) (https://digital.nhs.uk/about-nhs-digital/corporate-information-and-documents/independent-group-advising-on-the-release-of-data) via an application made in the Data Access Request Service (DARS) Online system (ref. DARS-NIC-381078-Y9C5K) (https://digital.nhs.uk/services/data-access-request-service-dars/dars-products-and-services). The CVD-COVID-UK/COVID-IMPACT Approvals & Oversight Board (https://bhfdatasciencecentre.org/areas/cvd-covid-uk-covid-impact/) subsequently granted approval to this project to access the data within NHS England’s SDE service for England. The de-identified data used in this study were made available to accredited researchers only. Those wishing to gain access to the data should contact in the first instance.

## Ethical approval

The North East – Newcastle and North Tyneside 2 research ethics committee provided ethical approval for the CVD-COVID-UK/COVID-IMPACT research programme (REC No 20/NE/0161) to access, within secure trusted research environments, unconsented, whole-population, de-identified data from electronic health records collected as part of patients’ routine healthcare.

## Supplementary Material

### Methods S1: SNOMED code lists for mental disorders

The General Practice Extraction Service Data for Pandemic Planning and Research (GDPPR) dataset includes a subset of all SNOMED codes used in the United Kingdom. The subset of codes included is defined by the primary care domain (PCD) reference set.^1^ The PCD reference set is also used by the Quality and Outcomes Framework (QOF) and current and historical versions of the reference set are available from NHS England.^2^ The available SNOMED codes are grouped into code clusters.^1^

To develop our SNOMED code lists for mental disorders we extracted codes from all of the clusters related to relevant mental disorder diagnoses: DEPR_COD (depression diagnosis codes), DEPRES_COD (depression resolved codes) and MH_COD (psychosis and schizophrenia and bipolar affective disease codes) from version 47.1 of the QOF cluster list (which was the most recent version at the time of developing the SMI code lists).^3^ Additionally, we mapped existing Read v2 and Clinical Terms Version 3 code lists^4^ for schizophrenia, bipolar disorder and depression to SNOMED, and identified codes available within the GDPPR dataset. We collated all SNOMED codes identified from the clusters, and from the mapping process and then identified codes for each of schizophrenia, bipolar disorder or depression.

All of our SNOMED code lists are available via the HDR UK Phenotype Library:

- **Schizophrenia:** https://phenotypes.healthdatagateway.org/phenotypes/PH1718
- **Bipolar disorder:** https://phenotypes.healthdatagateway.org/phenotypes/PH1719
- **Depression:** https://phenotypes.healthdatagateway.org/phenotypes/PH1720

## Methods S2: Additional description of covariates

### Admission timing

We used the MINAP hospital arrival time variable to determine whether the admission was during the day (6am – 6pm) or overnight.

### Admission day

We used the MINAP hospital arrival time variable to determine whether the admission was on a weekday or weekend.

### 4-month period of admission

We used the MINAP hospital arrival time variable to identify the 4-month period during which the MI admission occurred, starting from the pre-pandemic period of November 2019 to February 2020.

### Hospital

We ascertained the hospital the patient was admitted to by linking the MINAP record to HES APC records.

### Comorbid angina

We identified comorbid angina if any of the following occurred:

- The MINAP previous angina field (field 2.06) recorded “1. Yes”
- The patient had a diagnosis of angina recorded in GDPPR prior to their date of MI admission.
- The patient had a diagnosis of angina recorded in HES APC prior to their date of MI admission.

We identified diagnoses using existing SNOMED (GDPPR) and ICD-10 (HES APC) code lists for angina which are available on the HDR UK Phenotype Library.^1^

### Comorbid cerebrovascular disease

We identified comorbid cerebrovascular disease if any of the following occurred:

- The MINAP cerebrovascular disease field (field 2.10) recorded “1. Yes”
- The patient had a diagnosis of cerebrovascular disease recorded in GDPPR prior to their date of MI admission. We identified diagnoses using SNOMED codes in the haemorrhagic stroke (HSTRK_COD), non-haemorrhagic stroke (OSTR_COD), stroke diagnosis (STRK_COD) and transient ischaemic attack (TIA_COD) code clusters (see Methods S2 for further explanation of code clusters) and additionally the SNOMED codes 195163003, 67992007 and 73192008.
- The patient had a diagnosis of cerebrovascular disease recorded in HES APC prior to their date of MI admission. We identified diagnoses using ICD-10 codes I60-I69 and G45.

Our code lists for comorbid cerebrovascular disease are available on our GitHub repository (https://github.com/BHFDSC/CCU046_01) and via the HDR UK Phenotype Library: https://phenotypes.healthdatagateway.org/phenotypes/PH1791.

### Comorbid chronic renal failure

We identified comorbid chronic renal failure if any of the following occurred:

- The MINAP chronic renal failure field (field 2.12) recorded “1. Yes”
- The patient had a diagnosis of chronic renal failure recorded in GDPPR prior to their date of MI admission. We identified diagnoses using SNOMED codes. We developed the code list by identifying chronic renal failure codes from an existing chronic kidney disease code list^2^ and the GDPPR code cluster for chronic kidney disease stage 4 and 5 (CKDATRISK1_COD).
- The patient had a diagnosis of chronic renal failure recorded in HES APC prior to their date of MI admission. We identified diagnoses using ICD-10 codes. We developed the code list by identifying chronic renal failure codes from an existing chronic kidney disease code list^2^ and existing end stage renal disease code lists.^3,4^

Our code lists for comorbid chronic renal failure are available on our GitHub repository (https://github.com/BHFDSC/CCU046_01) and via the HDR UK Phenotype Library: https://phenotypes.healthdatagateway.org/phenotypes/PH1792.

### Comorbid diabetes

We identified comorbid diabetes if any of the following occurred:

- The MINAP diabetes field (field 2.17) recorded “1. Diabetes (dietary control)” or “2. Diabetes (oral medicine)” or “3. Diabetes (insulin)” or “5. Insulin plus oral medication”
- The patient had a diagnosis of diabetes recorded in GDPPR prior to their date of MI admission.
- The patient had a diagnosis of diabetes recorded in HES APC prior to their date of MI admission.

We identified diagnoses using existing SNOMED (GDPPR) and ICD-10 (HES APC) code lists for diabetes which are available on the HDR UK Phenotype Library.^5^

### Comorbid heart failure

We identified comorbid heart failure if any of the following occurred:

- The MINAP heart failure field (field 2.13) recorded “1. Yes”
- The patient had a diagnosis of heart failure recorded in GDPPR prior to their date of MI admission.
- The patient had a diagnosis of heart failure recorded in HES APC prior to their date of MI admission.

We identified diagnoses using existing SNOMED (GDPPR) and ICD-10 (HES APC) code lists for heart failure which are available on the HDR UK Phenotype Library.^6^

### Previous MI

We identified previous MI if either of the following occurred:

- The patient had a diagnosis of MI recorded in GDPPR more than 3 days prior to their date of MI admission (to ensure that we didn’t count the index event). We identified diagnoses by adapting an existing MI SNOMED code list.^7^
- The patient had a diagnosis of MI recorded in HES APC more than 3 days prior to their date of MI admission. We identified diagnoses using ICD-10 codes (I21, I22) as per previous studies from the authors.^8^

Our code lists for myocardial infarction are available on our GitHub repository (https://github.com/BHFDSC/CCU046_01) and via the HDR UK Phenotype Library (https://phenotypes.healthdatagateway.org/phenotypes/PH1722).

We couldn’t use the MINAP previous MI field (field 2.05) because it wasn’t included in the version of the MINAP dataset held by the CVD-COVID-UK/COVID-IMPACT Consortium at the time of data curation.

### Previous PCI

We identified previous PCI if any of the following occurred:

- The MINAP previous PCI field (field 2.18) recorded “1. Yes”
- The patient had a PCI recorded in HES APC prior to their date of MI admission. We identified PCI using the OPCS-4 procedure code K75, in line with previous studies from the authors.^8^

We didn’t use GDPPR to identify PCI because the GDPPR clusters do not include a comprehensive set of PCI codes.

### COVID-19 positive

We identified patients who had COVID-19 within 14 days either side of their MI admission based on diagnoses recorded in GDPPR or HES APC, hospitalised cases of COVID-19 recorded in CHESS and positive tests for COVID-19 recorded in SGSS or SUS.

### Guideline informed care standards

#### Receipt of angiography (NSTEMI)

We categorised receipt of angiography as:

- Received within 72 hours
- Received out with 72 hours
- Received, timing unknown
- Eligible but did not receive
- Not eligible

We derived this variable from the MINAP fields for coronary angiography (field 4.13), time of arrival at hospital (field 3.06) and time of local angiography (field 4.18).

#### Admission to cardiac ward (NSTEMI)

Admission to cardiac ward was included as a binary variable, defined as ‘yes’ if the MINAP admission ward field (field 3.17) was recorded as ‘cardiac care unit’, ‘intensive therapy unit’ or ‘cardiac ward (non CCU)’ and otherwise ‘no’ if the field was not recorded as unknown or left blank.

#### Receipt of primary PCI (STEMI)

We defined a binary receipt of PCI variable from the MINAP field initial reperfusion treatment (field 3.39).

#### Call-to-balloon (CTB) time within target (STEMI)

The MINAP report defines two targets for CTB times: 120 minutes and 150 minutes. We categorised CTB time as:

- Less than 120 minutes
- Between 120 and 150 minutes
- Greater than or equal to 150 mins
- Received pPCI but CTB time unknown
- Did not receive pPCI

We derived this variable from the MINAP fields initial reperfusion treatment (field 3.39), time of call for help (field 3.02) and time of reperfusion treatement (field 3.09).

#### Indicated secondary prevention medication prescribed at discharge (both types of MI)

Indicated secondary prevention medication prescribed at discharge was included as a binary variable, defined as ‘yes’ if where indicated, the patient was prescribed (or refused) aspirin (field 4.08), beta blocker (field 4.05), statin (field 4.07), either ACE inhibitor or angiotensin receptor antagonist (field 4.06), and either thienopyridine (field 4.27) or ticagrelor (field 4.31).

#### Referral for cardiac rehabilitation (both types of MI)

We categorised referral for cardiac rehabilitation as follows, based on the MINAP cardiac rehabilitation field (field 4.09)

- Yes
- No
- Not indicated
- Patient declined

---

**Figure S1:**
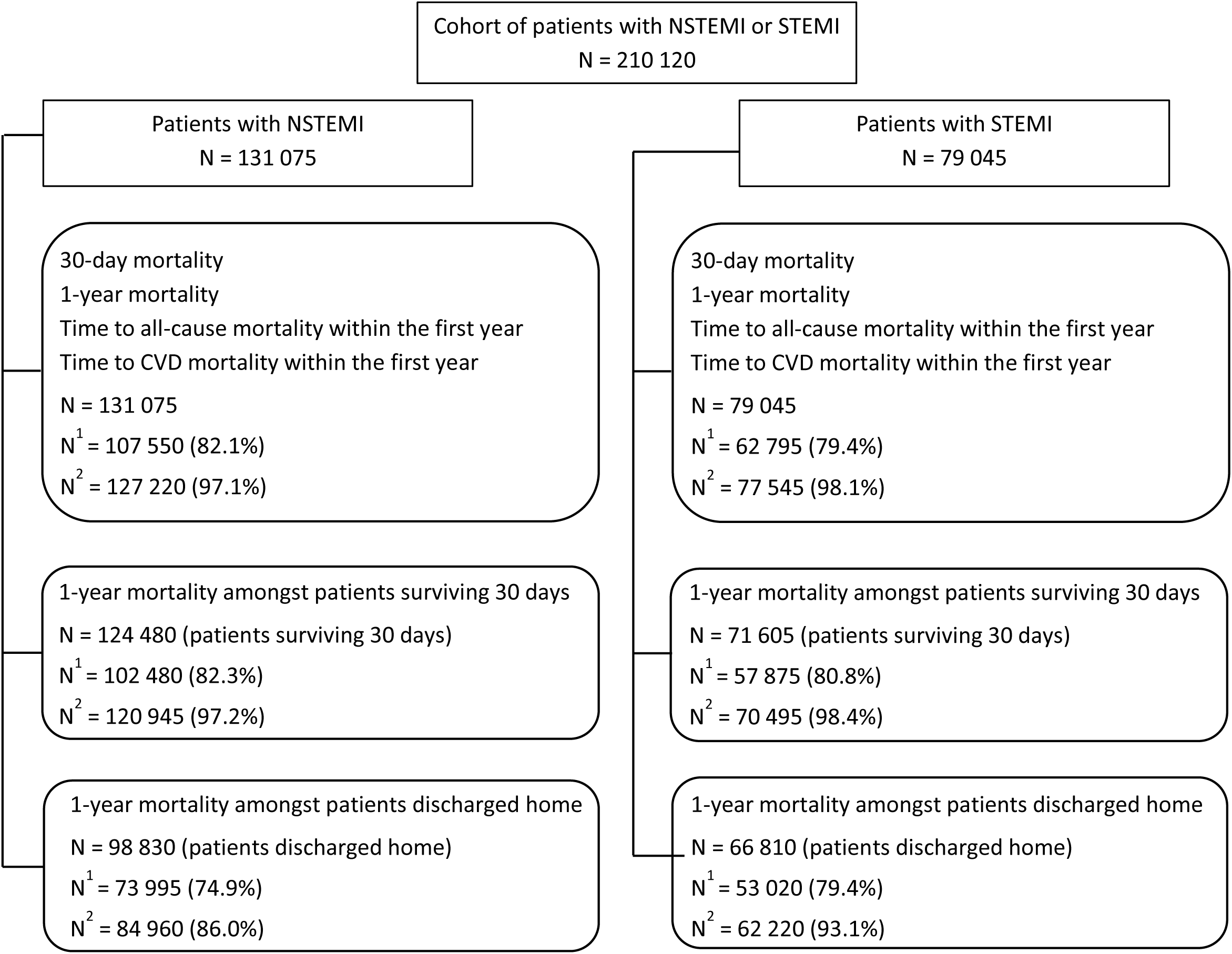
Flow diagram for the analysis cohorts. N = patients eligible for inclusion in analysis N^1^ = complete cases (with percentage of N) N^2^ = complete cases (sensitivity analysis) (with percentage of N) As per NHSE’s SDE statistical disclosure control rules, counts are rounded to the nearest 5 and counts less than 10 are suppressed

**Figure S2:**
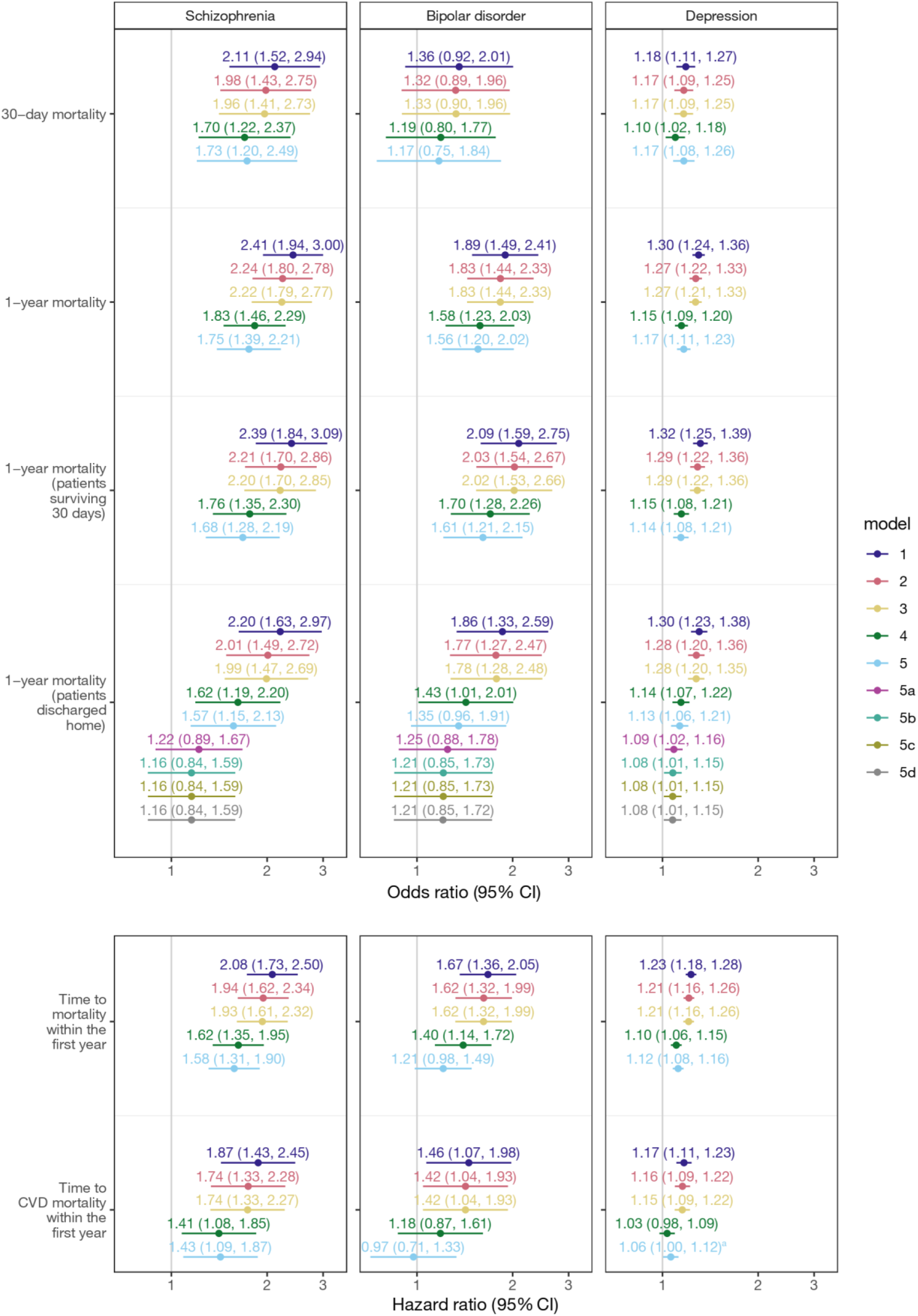
Odds ratios and hazard ratios for mortality following NSTEMI in patients with schizophrenia, bipolar disorder or depression versus those without any of these disorders. Models 1 to 5. Model 1 adjusts for age, sex and hospital (random effect); model 2 additionally adjusts for ethnicity and area-level deprivation; model 3 adds timing variables; model 4 adds clinical history; model 5 adds MI presentation features; model 5a adds receipt of angiography; 5b adds referral for cardiac rehabilitation; 5c adds indicated secondary prevention medication prescribed at discharge and 5d adds admission to cardiac ward. ^a^1.062 (1.004, 1.123)

**Figure S3:**
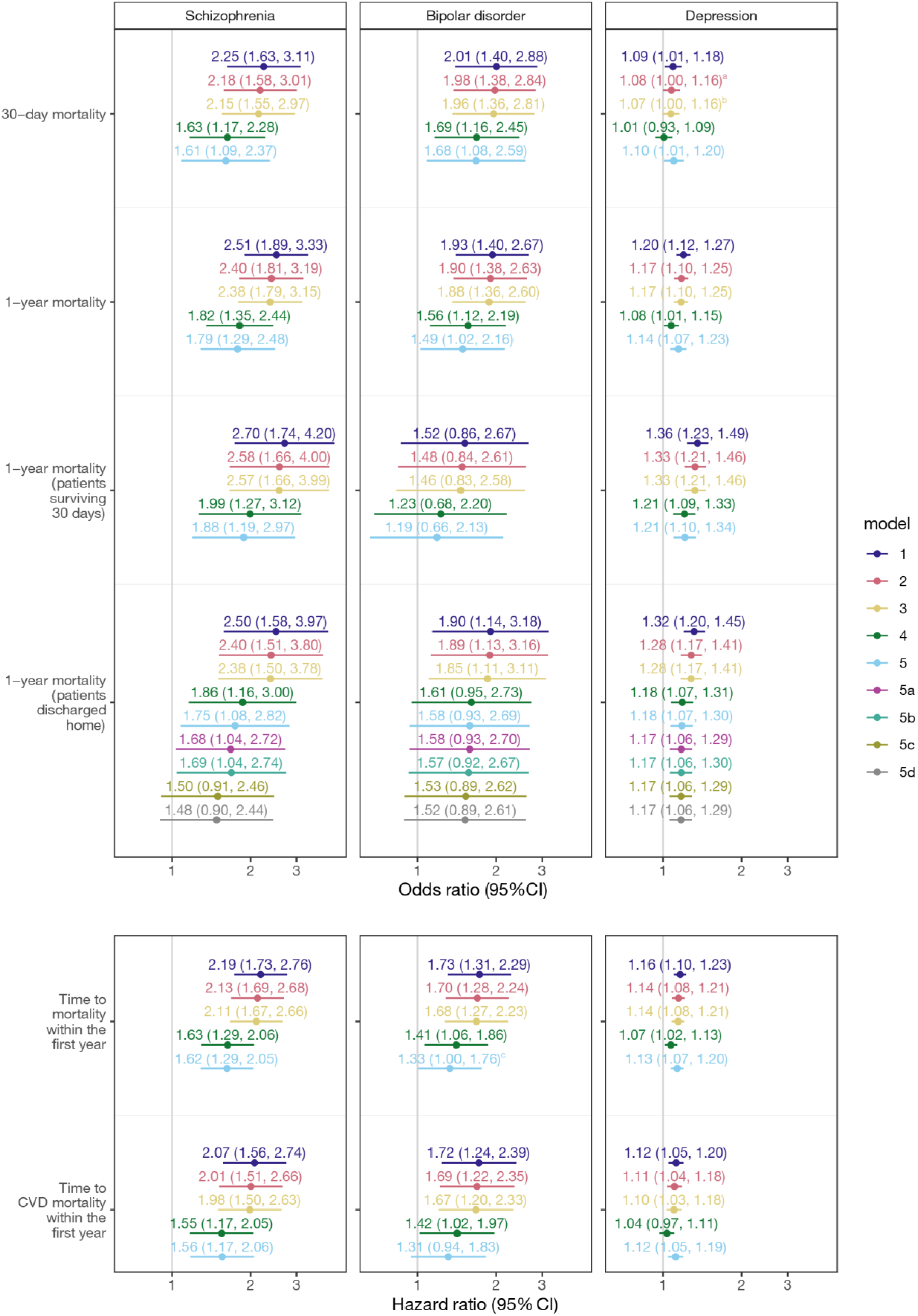
Odds ratios and hazard ratios for mortality following STEMI in patients with schizophrenia, bipolar disorder or depression versus those without any of these disorders. Models 1 to 5. Model 1 adjusts for age, sex and hospital (random effect); model 2 additionally adjusts for ethnicity and area-level deprivation; model 3 adds timing variables; model 4 adds clinical history; model 5 adds MI presentation features; model 5a adds receipt of primary PCI (yes/no); in model 5b the binary primary PCI is replaced by the categorical CTB time within target variable (<120 mins, 120–150 mins, ≥150 mins, received PCI but time unknown, did not receive PCI); 5c adds referral for cardiac rehabilitation and 5d adds indicated secondary prevention medication prescribed at discharge. ^a^1.078 (1.001, 1.161); ^b^1.073 (0.996, 1.155); ^c^1.331 (1.005, 1.762)

**Figure S4:**
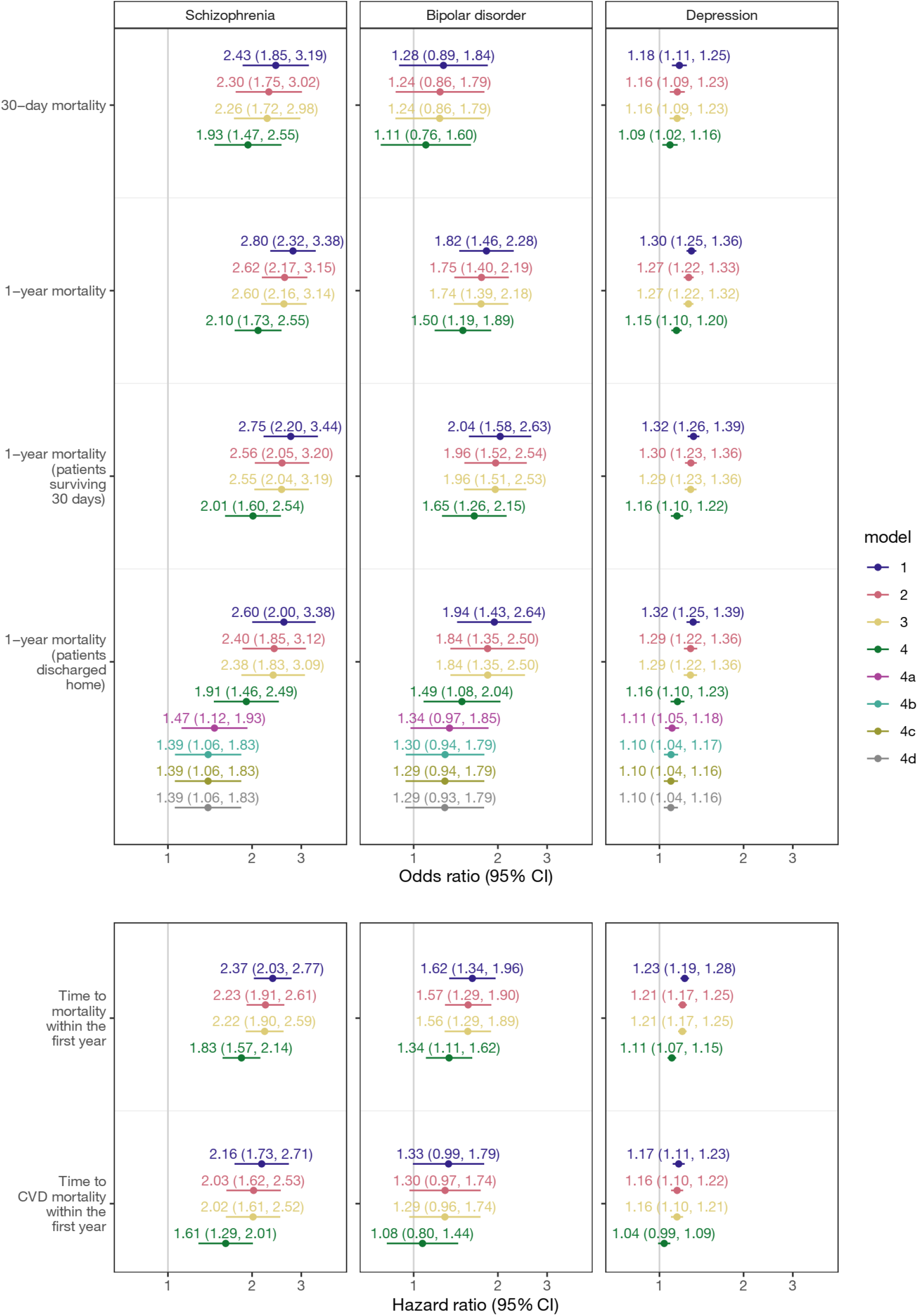
Odds ratios for receipt of each care standard following NSTEMI in patients with schizophrenia, bipolar disorder or depression versus those without any of these disorders. Sensitivity analysis. Models 1 to 4. Model 1 adjusts for age, sex and hospital (random effect); model 2 additionally adjusts for ethnicity and area-level deprivation; model 3 adds timing variables; model 4 adds clinical history; model 4a adds receipt of angiography; 4b adds referral for cardiac rehabilitation; 4c adds indicated secondary prevention medication prescribed at discharge and 4d adds admission to cardiac ward.

**Figure S5:**
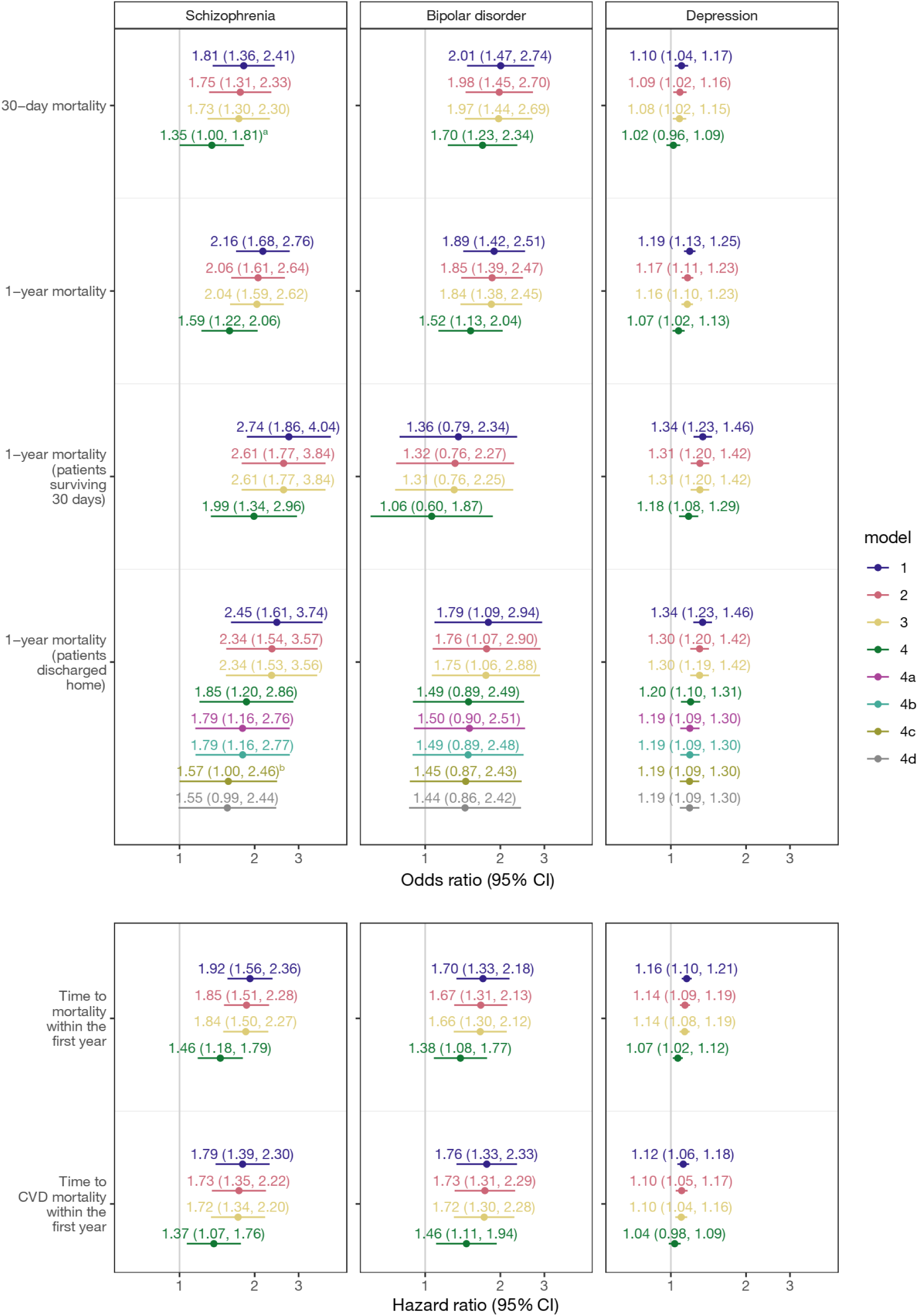
Odds ratios for receipt of each care standard following STEMI in patients with schizophrenia, bipolar disorder or depression versus those without any of these disorders. Sensitivity analysis. Models 1 to 4. Model 1 adjusts for age, sex and hospital (random effect); model 2 additionally adjusts for ethnicity and area-level deprivation; model 3 adds timing variables; model 4 adds clinical history; model 4a adds receipt of primary PCI (yes/no); in model 4b the binary primary PCI is replaced by the categorical CTB time within target variable (<120 mins, 120–150 mins, ≥150 mins, received PCI but time unknown, did not receive PCI); 4c adds referral for cardiac rehabilitation and 4d adds indicated secondary prevention medication prescribed at discharge. ^a^1.348 (1.002, 1.812), ^b^1.5685 (1.0001, 2.4598)

**Figure S6:**
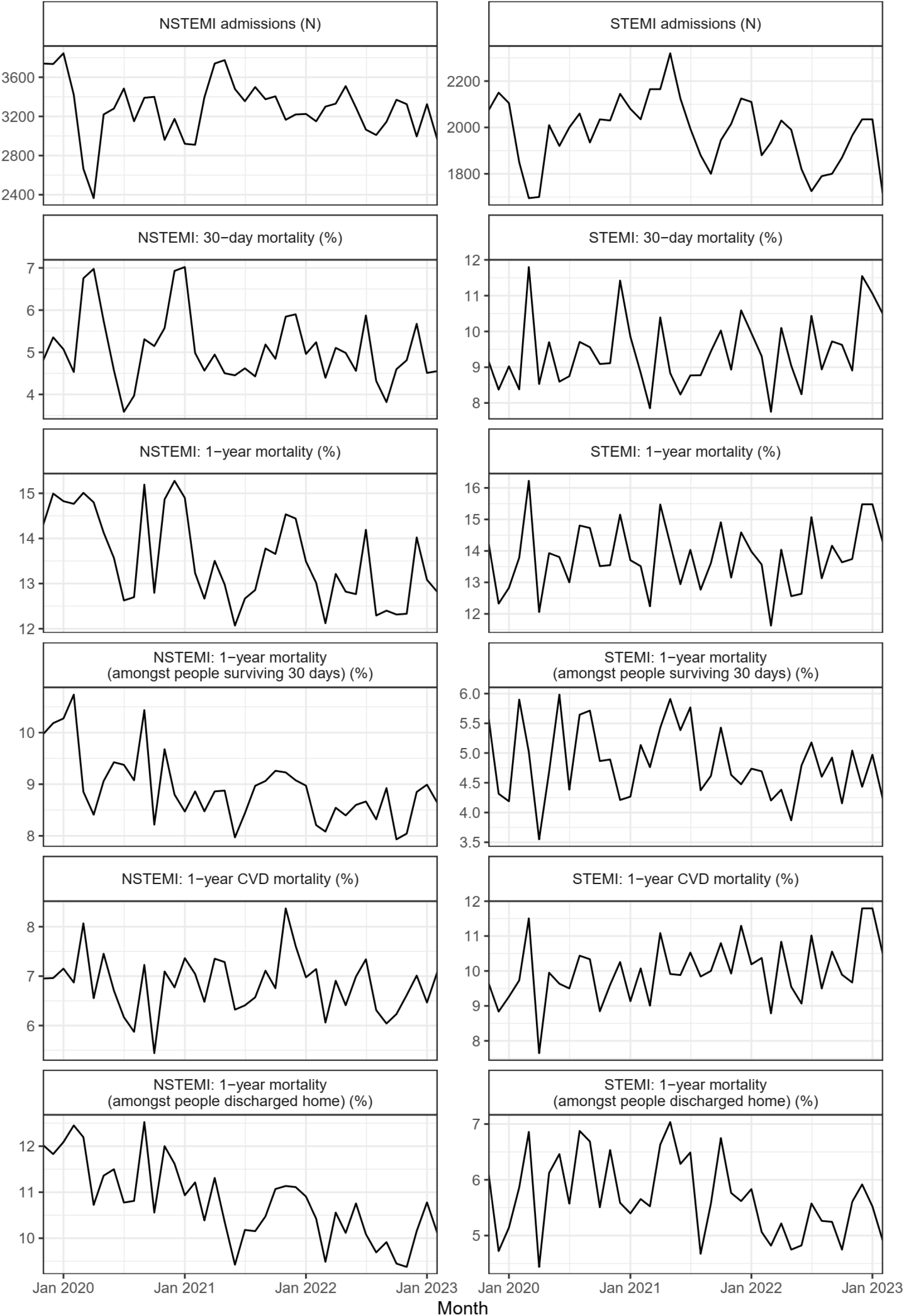
MI admissions and mortality by admission month.

**Figure S7:**
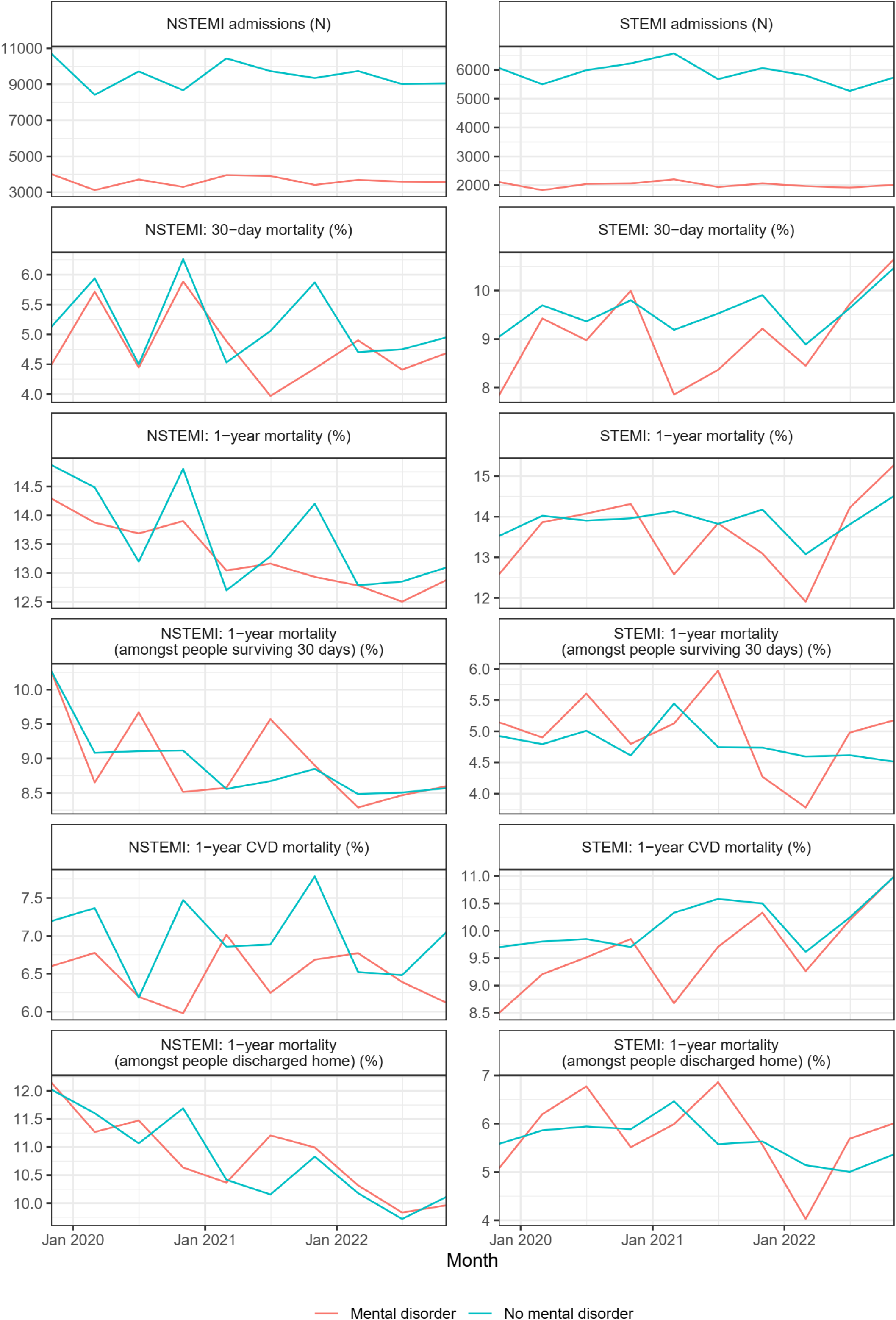
MI admissions and mortality by 4-month admission period and mental disorder.

**Table S1:**
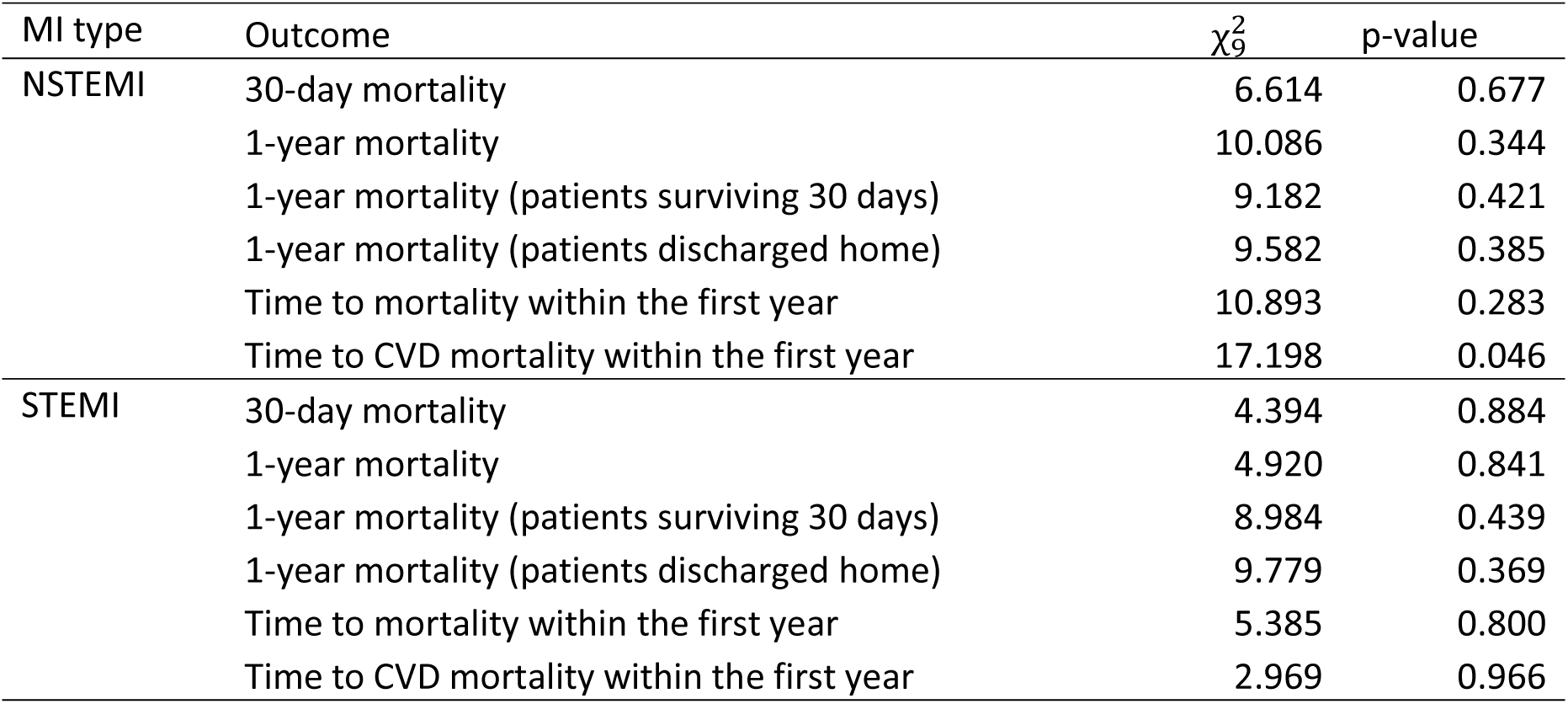
Interaction between mental disorder and 4-month time period; likelihood ratio tests of whether adding the interaction improved the fit of the model (primary analysis)

**Table S2:** Hazard ratios by 4-month time period for time to CVD mortality within the first year following NSTEMI in patients with versus without any of schizophrenia, bipolar disorder or depression. Model 5 with interaction between mental disorder and 4-month time period.

| 4-month time period | Hazard ratio (95% confidence interval) |
| --- | --- |
| Nov 2019 - Feb 2020 | 1.15 (0.99, 1.35) |
| Mar 2020 - Jun 2020 | 1.05 (0.88, 1.26) |
| Jul 2020 - Oct 2020 | 1.16 (0.98, 1.38) |
| Nov 2020 - Feb 2021 | 0.83 (0.69, 0.99) |
| Mar 2021 - Jun 2021 | 1.10 (0.94, 1.28) |
| Jul 2021 - Oct 2021 | 1.10 (0.93, 1.30) |
| Nov 2021 - Feb 2022 | 0.95 (0.80, 1.12) |
| Mar 2022 - Jun 2022 | 1.27 (1.07, 1.51) |
| Jul 2022 - Oct 2022 | 1.14 (0.95, 1.36) |
| Nov 2022 - Feb 2023 | 0.98 (0.82, 1.17) |

**Table S3:** Interaction between mental disorder and 4-month time period; likelihood ratio tests of whether adding the interaction improved the fit of the sensitivity analysis models.

| MI type | Outcome | $\chi^2_9$ | p-value |
| --- | --- | --- | --- |
| NSTEMI | 30-day mortality | 12.772 | 0.173 |
|  | 1-year mortality | 10.205 | 0.334 |
|  | 1-year mortality (patients surviving 30 days) | 9.091 | 0.429 |
|  | 1-year mortality (patients discharged home) | 9.727 | 0.373 |
|  | Time to mortality within the first year | 11.230 | 0.260 |
|  | Time to CVD mortality within the first year | 17.547 | 0.041 |
| STEMI | 30-day mortality | 5.710 | 0.769 |
|  | 1-year mortality | 5.323 | 0.805 |
|  | 1-year mortality (patients surviving 30 days) | 8.278 | 0.506 |
|  | 1-year mortality (patients discharged home) | 11.954 | 0.216 |
|  | Time to mortality within the first year | 4.841 | 0.848 |
|  | Time to CVD mortality within the first year | 4.956 | 0.838 |

**Table S4:** Hazard ratios by 4-month time period for time to CVD mortality within the first year following NSTEMI in patients with versus without any of schizophrenia, bipolar disorder or depression. Model 4 with interaction between mental disorder and 4-month time period. Sensitivity analysis.

| 4-month time period | Hazard ratio (95% confidence interval) |
| --- | --- |
| Nov 2019 - Feb 2020 | 1.07 (0.93, 1.24) |
| Mar 2020 - Jun 2020 | 1.01 (0.86, 1.18) |
| Jul 2020 - Oct 2020 | 1.17 (1.00, 1.36) |
| Nov 2020 - Feb 2021 | 0.85 (0.72, 1.00) |
| Mar 2021 - Jun 2021 | 1.16 (1.00, 1.33) |
| Jul 2021 - Oct 2021 | 1.04 (0.90, 1.21) |
| Nov 2021 - Feb 2022 | 0.96 (0.83, 1.12) |
| Mar 2022 - Jun 2022 | 1.21 (1.04, 1.40) |
| Jul 2022 - Oct 2022 | 1.15 (0.98, 1.34) |
| Nov 2022 - Feb 2023 | 0.96 (0.82, 1.12) |

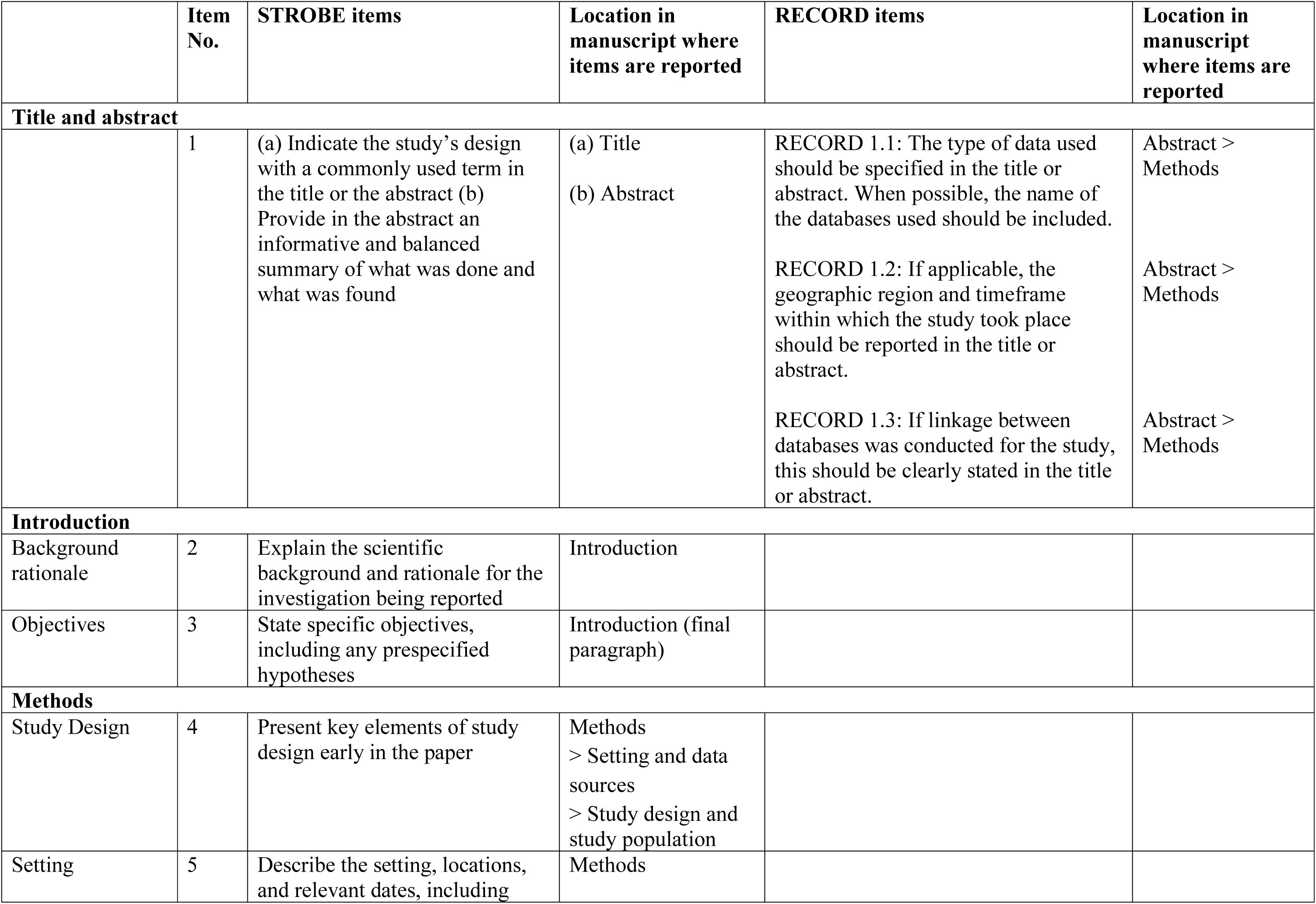
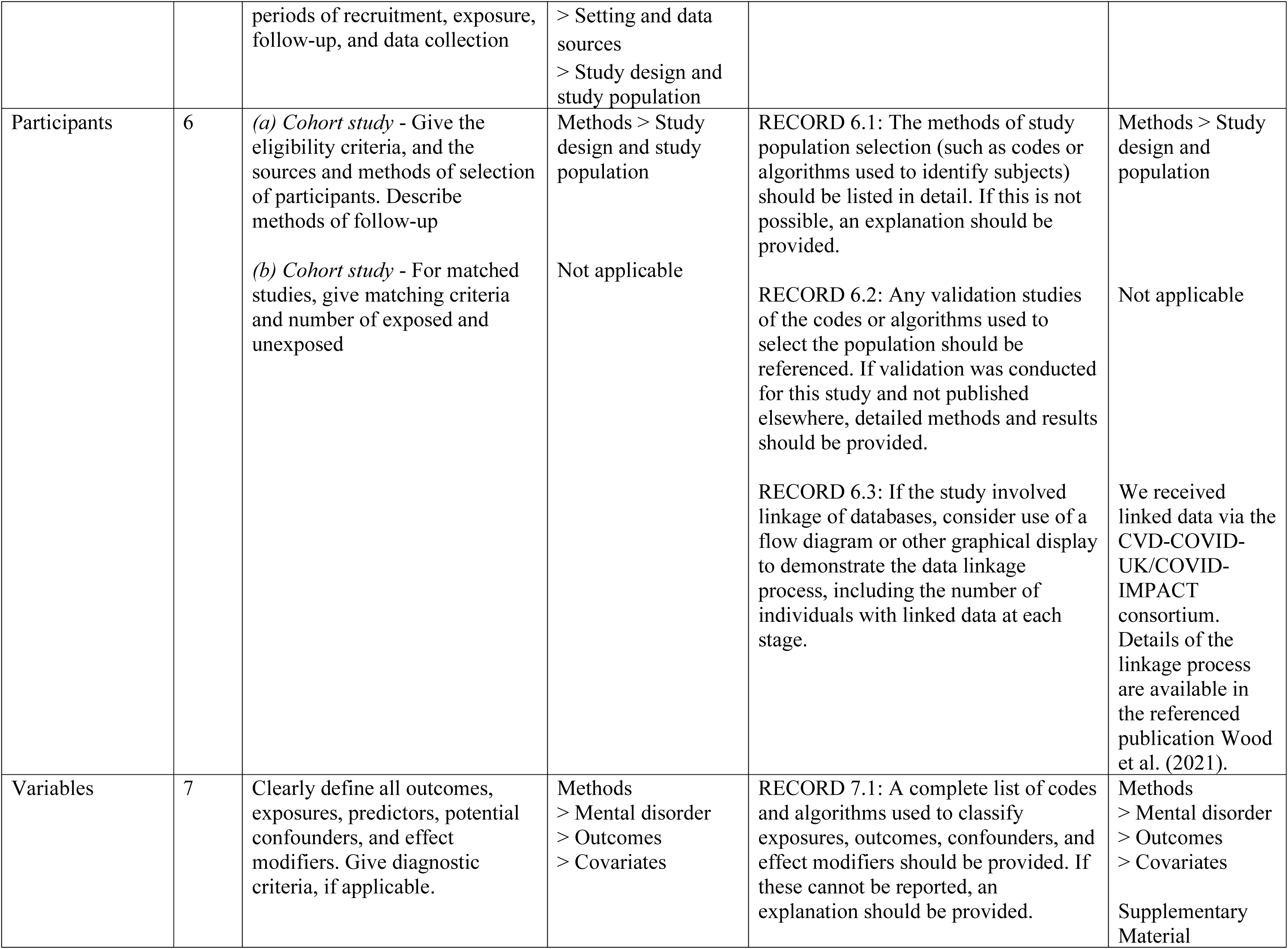
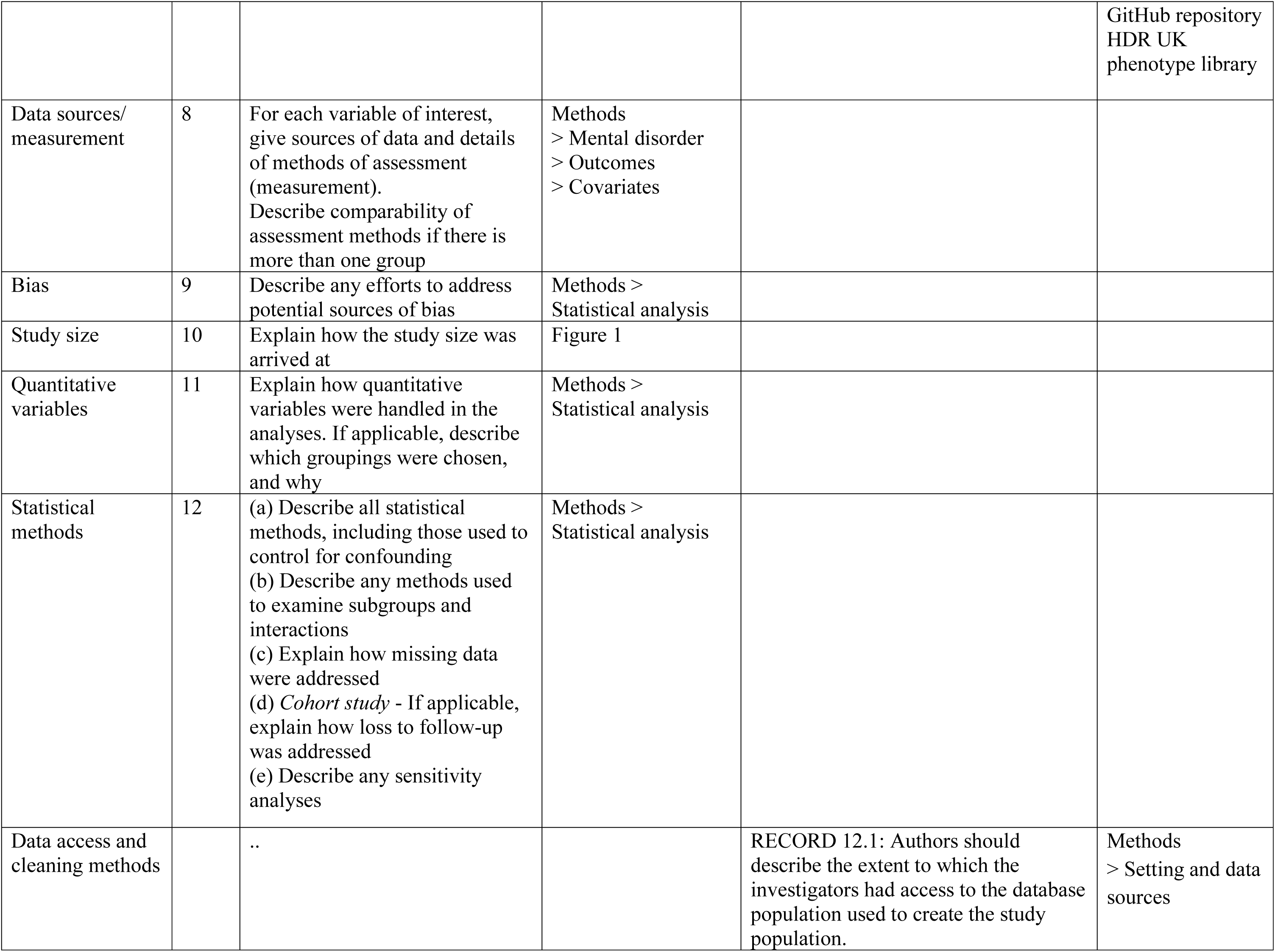
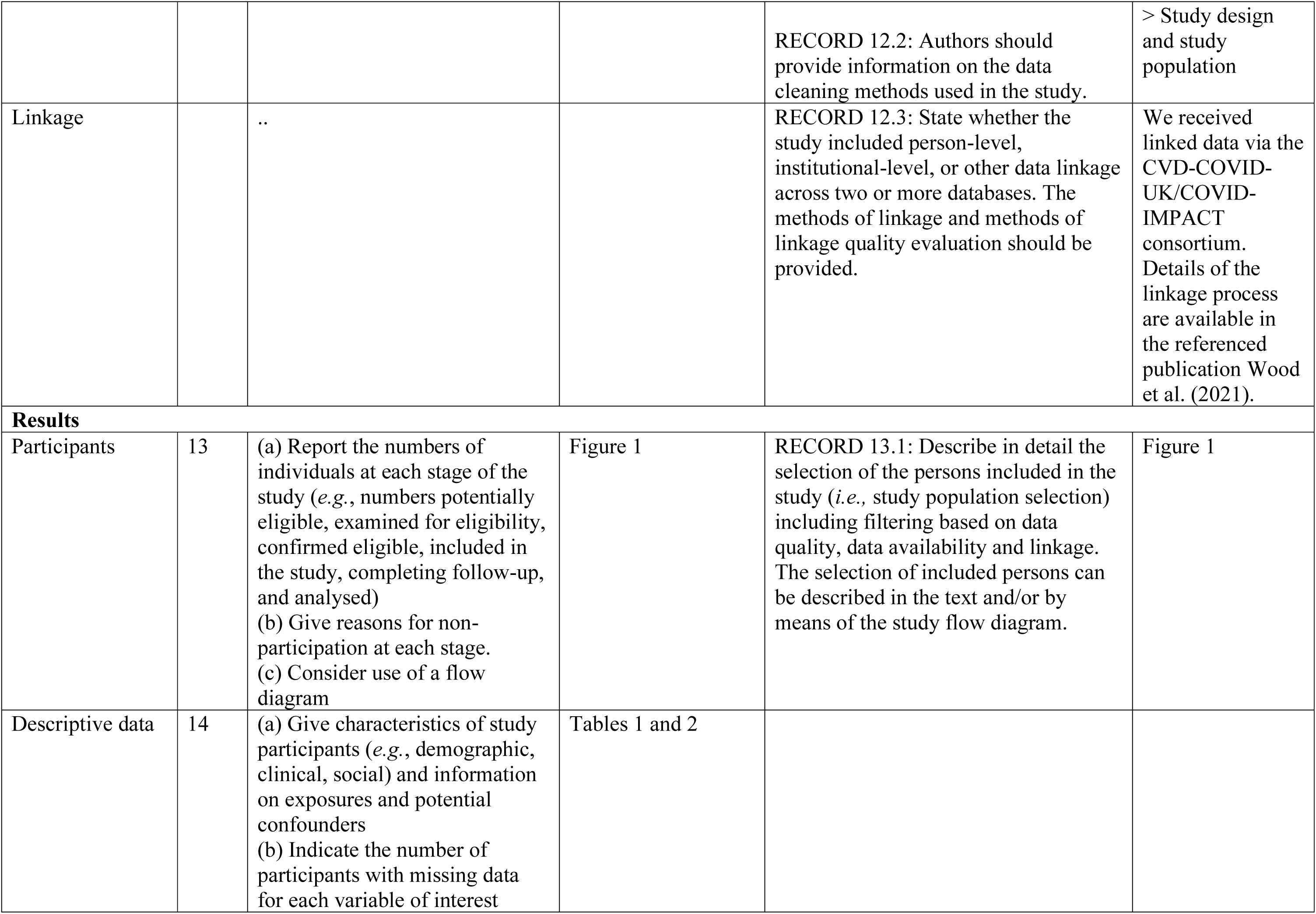
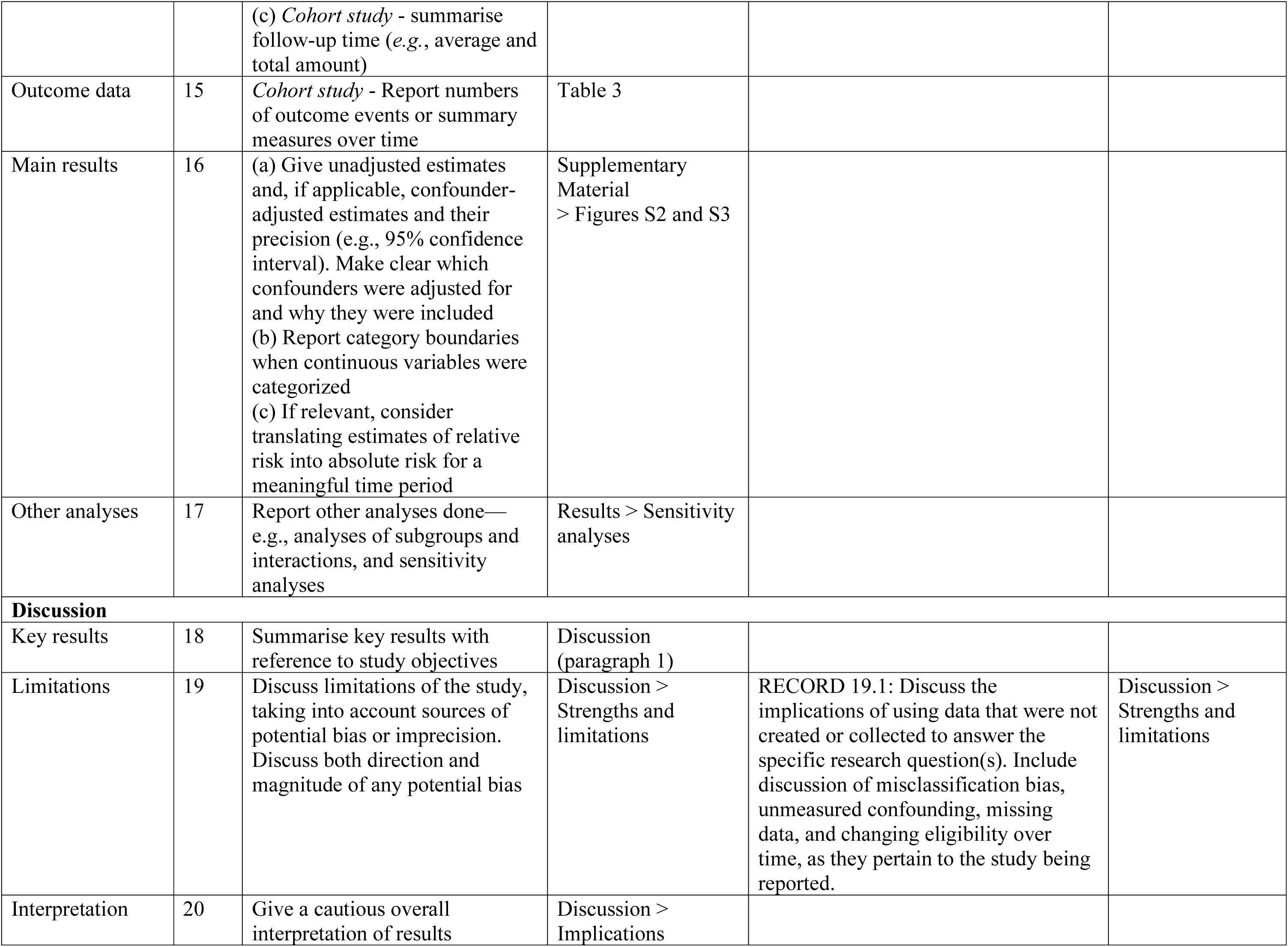
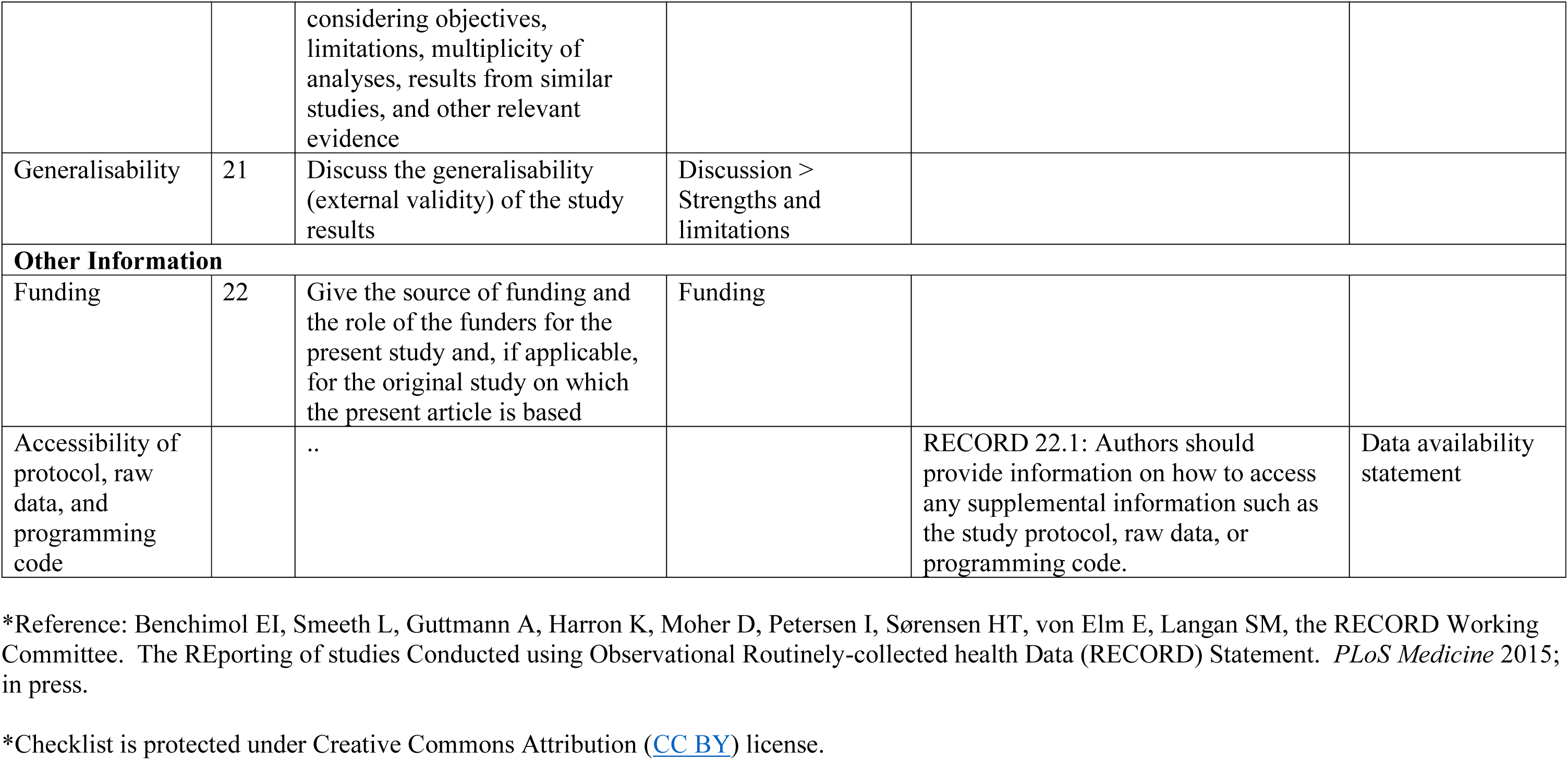
The RECORD statement – checklist of items, extended from the STROBE statement, that should be reported in observational studies using routinely collected health data.

|  | Item No. | STROBE items | Location in manuscript where items are reported | RECORD items | Location in manuscript where items are reported |
| --- | --- | --- | --- | --- | --- |
| **Title and abstract** |  |  |  |  |  |
|  | 1 | (a) Indicate the study's design with a commonly used term in the title or the abstract (b) Provide in the abstract an informative and balanced summary of what was done and what was found | (a) Title<br>(b) Abstract | RECORD 1.1: The type of data used should be specified in the title or abstract. When possible, the name of the databases used should be included.<br>RECORD 1.2: If applicable, the geographic region and timeframe within which the study took place should be reported in the title or abstract.<br>RECORD 1.3: If linkage between databases was conducted for the study, this should be clearly stated in the title or abstract. | Abstract > Methods<br>Abstract > Methods<br>Abstract > Methods |
| **Introduction** |  |  |  |  |  |
| Background rationale | 2 | Explain the scientific background and rationale for the investigation being reported | Introduction |  |  |
| Objectives | 3 | State specific objectives, including any prespecified hypotheses | Introduction (final paragraph) |  |  |
| **Methods** |  |  |  |  |  |
| Study Design | 4 | Present key elements of study design early in the paper | Methods<br>> Setting and data sources<br>> Study design and study population |  |  |
| Setting | 5 | Describe the setting, locations, and relevant dates, including | Methods |  |  |
|  |  | periods of recruitment, exposure, follow-up, and data collection | > Setting and data sources<br>> Study design and study population |  |  |
| Participants | 6 | (a) *Cohort study* - Give the eligibility criteria, and the sources and methods of selection of participants. Describe methods of follow-up<br>(b) *Cohort study* - For matched studies, give matching criteria and number of exposed and unexposed | Methods > Study design and study population<br>Not applicable | RECORD 6.1: The methods of study population selection (such as codes or algorithms used to identify subjects) should be listed in detail. If this is not possible, an explanation should be provided.<br>RECORD 6.2: Any validation studies of the codes or algorithms used to select the population should be referenced. If validation was conducted for this study and not published elsewhere, detailed methods and results should be provided.<br>RECORD 6.3: If the study involved linkage of databases, consider use of a flow diagram or other graphical display to demonstrate the data linkage process, including the number of individuals with linked data at each stage. | Methods > Study design and population<br>Not applicable<br>We received linked data via the CVD-COVID-UK/COVID-IMPACT consortium. Details of the linkage process are available in the referenced publication Wood et al. (2021). |
| Variables | 7 | Clearly define all outcomes, exposures, predictors, potential confounders, and effect modifiers. Give diagnostic criteria, if applicable. | Methods<br>> Mental disorder<br>> Outcomes<br>> Covariates | RECORD 7.1: A complete list of codes and algorithms used to classify exposures, outcomes, confounders, and effect modifiers should be provided. If these cannot be reported, an explanation should be provided. | Methods<br>> Mental disorder<br>> Outcomes<br>> Covariates<br>Supplementary Material |
|  |  |  |  |  | GitHub repository<br>HDR UK<br>phenotype library |
| Data sources/<br>measurement | 8 | For each variable of interest, give sources of data and details of methods of assessment (measurement).<br>Describe comparability of assessment methods if there is more than one group | Methods<br>> Mental disorder<br>> Outcomes<br>> Covariates |  |  |
| Bias | 9 | Describe any efforts to address potential sources of bias | Methods ><br>Statistical analysis |  |  |
| Study size | 10 | Explain how the study size was arrived at | Figure 1 |  |  |
| Quantitative variables | 11 | Explain how quantitative variables were handled in the analyses. If applicable, describe which groupings were chosen, and why | Methods ><br>Statistical analysis |  |  |
| Statistical methods | 12 | (a) Describe all statistical methods, including those used to control for confounding<br>(b) Describe any methods used to examine subgroups and interactions<br>(c) Explain how missing data were addressed<br>(d) *Cohort study* - If applicable, explain how loss to follow-up was addressed<br>(e) Describe any sensitivity analyses | Methods ><br>Statistical analysis |  |  |
| Data access and cleaning methods |  | .. |  | RECORD 12.1: Authors should describe the extent to which the investigators had access to the database population used to create the study population. | Methods<br>> Setting and data sources |
|  |  |  |  | RECORD 12.2: Authors should provide information on the data cleaning methods used in the study. | > Study design and study population |
| Linkage |  | .. |  | RECORD 12.3: State whether the study included person-level, institutional-level, or other data linkage across two or more databases. The methods of linkage and methods of linkage quality evaluation should be provided. | We received linked data via the CVD-COVID-UK/COVID-IMPACT consortium. Details of the linkage process are available in the referenced publication Wood et al. (2021). |
| **Results** |  |  |  |  |  |
| Participants | 13 | (a) Report the numbers of individuals at each stage of the study (*e.g.*, numbers potentially eligible, examined for eligibility, confirmed eligible, included in the study, completing follow-up, and analysed)<br>(b) Give reasons for non-participation at each stage.<br>(c) Consider use of a flow diagram | Figure 1 | RECORD 13.1: Describe in detail the selection of the persons included in the study (*i.e.*, study population selection) including filtering based on data quality, data availability and linkage. The selection of included persons can be described in the text and/or by means of the study flow diagram. | Figure 1 |
| Descriptive data | 14 | (a) Give characteristics of study participants (*e.g.*, demographic, clinical, social) and information on exposures and potential confounders<br>(b) Indicate the number of participants with missing data for each variable of interest (c) *Cohort study* - summarise follow-up time (e.g., average and total amount) | Tables 1 and 2 |  |  |
| Outcome data | 15 | *Cohort study* - Report numbers of outcome events or summary measures over time | Table 3 |  |  |
| Main results | 16 | (a) Give unadjusted estimates and, if applicable, confounder-adjusted estimates and their precision (e.g., 95% confidence interval). Make clear which confounders were adjusted for and why they were included<br>(b) Report category boundaries when continuous variables were categorized<br>(c) If relevant, consider translating estimates of relative risk into absolute risk for a meaningful time period | Supplementary Material<br>> Figures S2 and S3 |  |  |
| Other analyses | 17 | Report other analyses done—e.g., analyses of subgroups and interactions, and sensitivity analyses | Results > Sensitivity analyses |  |  |
| **Discussion** |  |  |  |  |  |
| Key results | 18 | Summarise key results with reference to study objectives | Discussion (paragraph 1) |  |  |
| Limitations | 19 | Discuss limitations of the study, taking into account sources of potential bias or imprecision. Discuss both direction and magnitude of any potential bias | Discussion > Strengths and limitations | RECORD 19.1: Discuss the implications of using data that were not created or collected to answer the specific research question(s). Include discussion of misclassification bias, unmeasured confounding, missing data, and changing eligibility over time, as they pertain to the study being reported. | Discussion > Strengths and limitations |
| Interpretation | 20 | Give a cautious overall interpretation of results considering objectives, limitations, multiplicity of analyses, results from similar studies, and other relevant evidence | Discussion > Implications |  |  |
| Generalisability | 21 | Discuss the generalisability (external validity) of the study results | Discussion > Strengths and limitations |  |  |
| **Other Information** |  |  |  |  |  |
| Funding | 22 | Give the source of funding and the role of the funders for the present study and, if applicable, for the original study on which the present article is based | Funding |  |  |
| Accessibility of protocol, raw data, and programming code |  | .. |  | RECORD 22.1: Authors should provide information on how to access any supplemental information such as the study protocol, raw data, or programming code. | Data availability statement |

\*Reference: Benchimol EI, Smeeth L, Guttman A, Harron K, Moher D, Petersen I, Sørensen HT, von Elm E, Langan SM, the RECORD Working Committee. The REporting of studies Conducted using Observational Routinely-collected health Data (RECORD) Statement. *PLoS Medicine* 2015; in press.
\*Checklist is protected under Creative Commons Attribution ([CC BY](https://creativecommons.org/licenses/by/4.0/)) license.

